# Improving Access and Efficiency of Acute Ischemic Stroke Treatment Across Four Canadian Provinces: A Stepped-Wedge Trial

**DOI:** 10.1101/2024.09.13.24313666

**Authors:** Noreen Kamal, Elena A. Cora, Simone Alim, Judah Goldstien, David Volders, Shadi Aljendi, Heather Williams, Patrick T. Fok, Etienne van der Linde, Trish Helm-Neima, Renee Cashin, Brian Metcalfe, Julie Savoie BNSc, Wendy Simpkin, Cassie Chisholm, Michael D Hill, Bijoy K. Menon, Stephen Phillips, the ACTEAST Collaborators

**Affiliations:** Department of Industrial Engineering, Faculty of Engineering, Dalhousie University, Halifax, Canada; Department of Community Health and Epidemiology, Faculty of Medicine, Dalhousie University, Halifax, Canada; Department of Medicine (Division of Neurology), Faculty of Medicine, Dalhousie University, Halifax, Canada; Nova Scotia Health, Halifax, Canada; Department of Diagnostic Radiology, Faculty of Medicine, Dalhousie University, Halifax, Canada; Department of Mathematics and Statistics, Faculty of Science, Dalhousie University, Halifax, Canada; Department of Emergency Medicine, Faculty of Medicine, Dalhousie University, Halifax, Canada; Emergency Health Services, Halifax, Canada; EHS LifeFlight, Nova Scotia, Canada; Faculty of Computer Science, University of New Brunswick, Fredericton, Canada; Health PEI, Charlottetown, Canada; Dr. GB Cross Memorial Hospital, NL Health Services, Clarenville, Canada; Discipline of Emergency Medicine, Faculty of Medicine, Memorial University, St. John’s, Canada; Medflight NL, St. John’s, Canada; Vitalité Health Network, Moncton, Canada; NL Health Services, St. John’s, Canada; Department of Clinical Neurosciences, Radiology and Community Health Sciences, Cumming School of Medicine, University of Calgary, Calgary, Canada

## Abstract

**Background:** The translation of standard-of-care in acute ischemic stroke reperfusion interventions into practice is well established, but multifactorial obstacles exist in the complete adoption, which has led to inequities in access and delivery of services. The objective of this study was to improve access and efficiency of ischemic stroke treatment across four Atlantic Canadian Provinces.

**Methods:** A stepped-wedge cluster trial was conducted over 30 months with 3 clusters covering 34 sites. The trial was conducted across all 4 Atlantic Canadian provinces: Nova Scotia (NS), New Brunswick (NB), Prince Edward Island (PE), and Newfoundland and Labrador (NL). The design was quasi-randomized, with each cluster associated with one or more provinces: cluster 1 – NS; cluster 2 – NB and PE; and cluster 3 – NL. The patient population was all ischemic stroke patients across all 4 provinces. The intervention was a 6-month modified Quality Improvement Collaborative (mQIC), which was modified from the Breakthrough Series Collaborative to be half of the 1-year period and conducted virtually. The intervention consisted of assembling an interdisciplinary improvement team, 2 full-day workshops, webinars, and virtual site visits. Suggested changes included 6 process improvement strategies.

**Results:** Over the trial period, 8594 ischemic stroke patients were included, out of which 1576 patients received acute treatment. The proportion of patients that received treatment did not increase significantly with the intervention [0.4% increase for patients that received thrombolysis and/or EVT (p=0.68)]. Median door-to-needle time was reduced by 9.2 minutes with the intervention (p=0.01). Cluster 3 saw the greatest improvements in both access and efficiency.

**Conclusions:** A mQIC intervention resulted in improvement of process measures like door-to-needle time. Quality improvement initiatives may need to be longer to allow full implementation and tailored for each health system to ensure that each system sees improvement. In-person activities might be critical to ensure fidelity of the intervention.

## Introduction

Revolutionary advances in stroke treatment have occurred over the past two decades from thrombolysis using intravenous (IV) alteplase in the mid-1990’s^1,2,3^ and Tenecteplase^4^ to the most recent ground-breaking trials in 2015 showing the high efficacy of endovascular thrombectomy (EVT).^5,6^ These treatments can reduce disability due to stroke,^7^ thus reducing the need for assistance with daily living tasks,^8^ increase return to the workplace,^9^ and reduce hospital costs.^10^

Although treatment of acute ischemic stroke patients with alteplase and EVT are part of guideline care in Canada and around the world,^11,12^ less than optimal utilization rates for both of these treatments are observed. This lag in the translation of best standard of care is well known, which has led to increasing research in implementation science.^13^ In acute ischemic stroke treatment, the substantial economic and societal benefits of these therapies continue to make it critical to pursue optimal uptake. The lag in the uptake of best standard of care is exacerbated by in equities due to geographic, economic and demographic disparities, which has created a split between urban and rural access to treatment. Furthermore, access to EVT in rural areas is even more challenging because the treatment is relatively new and requires specialized equipment and personnel. Although both of these therapies mitigate disability, their effectiveness has been shown to be highly time dependent. In stroke treatment, minutes matter,^5,14^ bringing to light the mantra, “time is brain”.^15^ Therefore, there have been calls to reduce hospital thrombolysis treatment times to 30 minutes from hospital arrival,^10,16^ and to create seamless transfer processes for more efficient access to EVT for patients living outside major urban centers.^17,18^ Atlantic Canada is comprised of four provinces: Nova Scotia (NS), New Brunswick (NB), Prince Edward Island (PE), and Newfoundland and Labrador (NL). Stroke treatment in Atlantic Canada is particularly challenging because of small populations with a significant proportion that live in rural areas.

Using a stepped-wedge cluster trial design, this study [ACTEAST (Atlantic Canada Together Enhancing Acute Stroke Treatment)] aimed to evaluate if access to and efficiency of acute ischemic stroke treatment with iv thrombolysis and EVT across Atlantic Canada would improve with a *modified Quality Improvement Collaborative (mQIC) intervention*.^19, 20^ The objective of the cluster trial is to increase the proportion of ischemic stroke patients that receive reperfusion therapy and reduce the time to treatment.

## Methods

The ACTEAST Project was a 30-month stepped-wedge cluster trial from May 1 2020 to October 31 2022. The trial was quasi-randomized in-so-far as the sites were not randomized to a cluster for pragmatic reasons; rather, each cluster was associated with a province. A quasi-randomization methodology was used because of the close network of all stroke centers across each province, which would make it difficult to not contaminate each cluster if sites were to be randomized to different clusters. Each cluster had a single center that was capable of EVT treatment with the remainder of sites only capable of thrombolysis treatment; patients at these thrombolysis-only sites who were eligible for EVT were transferred to the EVT-capable center in their province. There were a total of 34 sites with 10 sites in cluster 1, 12 sites in cluster 2, and 12 sites in cluster 3. The stepped-wedge cluster trial design is shown in Figure 1. This trial took place entirely during the COVID-19 pandemic where restriction and additional processes were required including testing all patients for COVID-19. These restrictions also limited the administration of intervention activities to being entirely virtual.

**Figure 1:**
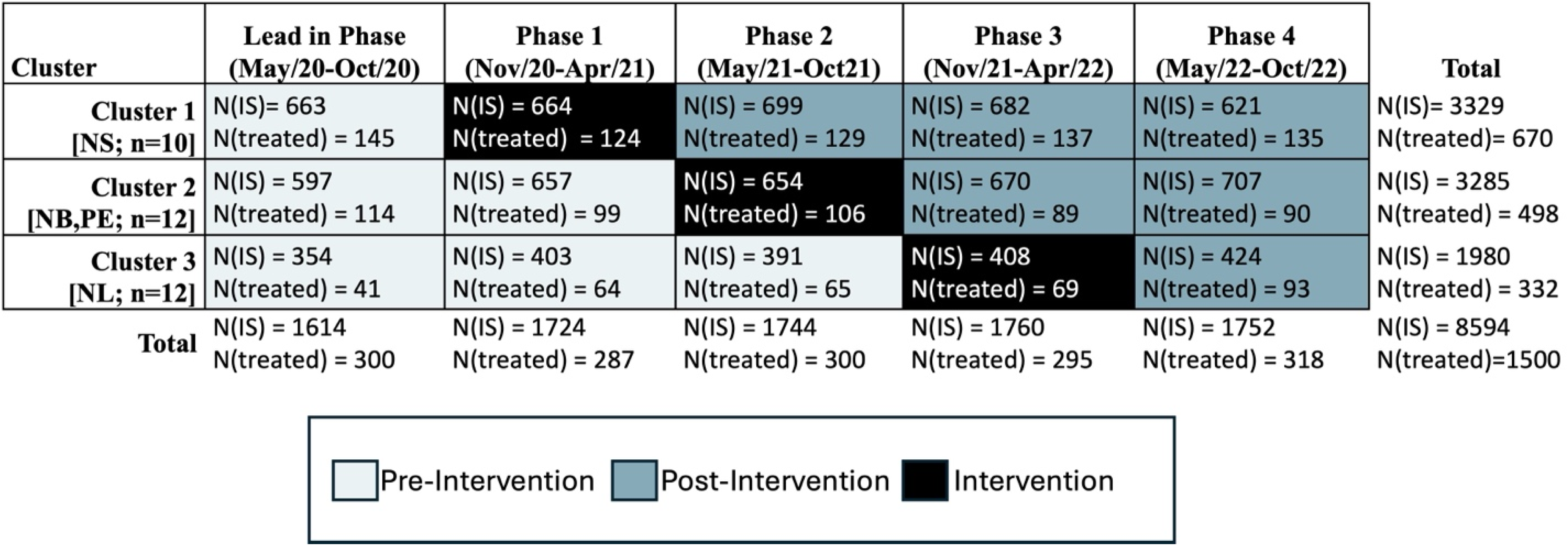
Stepped-wedge trial design and timeline. NS: Nova Scotia, NB: New Brunswick, PE: Prince Edward Island, NL: Newfoundland and Labrador; n denotes the number of sites in each cluster; N(IS): number of all ischemic stroke patients; N(treated): number of community onset ischemic stroke patients treated with thrombolysis and/or Endovascular Thrombectomy.

### Setting

ACTEAST was conducted across all four Atlantic Canadian provinces, which are smaller and less populated provinces as compared to the other Canadian provinces and have a combined population of approximately 2.5 million people. The Atlantic provinces have a larger percent of its population living in rural areas: in NS, 43% of its population live in rural areas, 48% in NB; 53% in PE; and 41% in NL. This is more than double other Canadian provinces; for example, only 14%, 17%, 14%, and 19% live in rural areas for Ontario, Alberta, British Columbia, and Quebec, respectively.^21^ Atlantic Canada also has an older population.^22^ The age and risk standardized stroke incidence rate is greater in Atlantic Canada than in other provinces: up to 140 strokes/100,000 people/year in the Atlantic provinces compared to 113 strokes/100,000 people/year in Ontario.^23^ All provinces have a provincially administered universal healthcare system. Each province has developed a provincial stroke system, where specific hospitals have been designated as stroke centers. These designated stroke centers have CT (computed tomography) scanners and expertise to treat patients with thrombolysis. They have access to stroke neurology expertise through telehealth. Ambulances bypass closer hospitals to take patients directly to a designated stroke center, which are distributed across each province to ensure optimal access for the population in each province.

EVT services were available in NS and NB in the cities of Halifax and Saint John respectively. PE has set-up cross-provincial transfer agreement with the EVT-capable hospital in Halifax, NS. NL did not have EVT services until June 20, 2022, which was completed during the trial period. Initially, only local patients had access to EVT, so there were no patients transferred for EVT in NL during the trial period. None of the provinces were using a pre-hospital LVO screening tool during the trial period, and all centers have access to CTA for determination of LVO. The utilization rates for ambulance use by acute ischemic stroke patients for each province is not specially known, but previous study found that 66% of all admitted stroke and TIA (transient ischemic attack) patients arrived by ambulance in Canada.^24^

### Site Enrolment and Patient Population

Enrolment into the ACTEAST trial was done at a site level. The Principal Investigator (NK) developed relationships with key medical and administrative personnel in all provinces to ensure that all designated stroke centers enrolled. All enrolled sites were required to set-up an interdisciplinary improvement team that consisted of paramedics, emergency department nurses, emergency physicians, stroke physicians (if applicable), stroke nurses (if applicable), CT technologists, radiologists, and administrators. Each team member was enrolled into the study and consented their participation.

### Measures

To measure access to treatment, the proportion of all ischemic stroke patients who received treatment was used. There were 3 separate measures for this: proportion that received iv thrombolysis only; proportion that received EVT only; and proportion that received iv thrombolysis and/or EVT (any treatment). We also assessed the proportion of patients that were transferred for EVT from a thrombolysis-only center.

To assess speed at which treatment was administered (treatment efficiency), all community onset ischemic stroke patients that received treatment were included and stroke that occurred in-hospital were excluded. Thrombolysis efficiency was assessed using door-to-needle time (time from hospital arrival to the start of thrombolysis treatment) and 911-to-needle time (time from 911 call to start of thrombolysis treatment). EVT efficiency was assessed using door-to-arterial-access time (time from hospital arrival to the start of EVT treatment) and 911-to-arterial-access time (time from 911 call to start of EVT treatment). Transfer efficiency was assessed using door-in-door-out time, which is the time from arrival at the thrombolysis-only center to departure from this center to the EVT-capable center.

Patient outcomes were measured using a pragmatic approach by utilizing hospital discharge disposition. The outcome measures included proportion of patients discharged home and proportion of patients that died in-hospital. This was done for all ischemic stroke patients and for all treated stroke patients (excluding in-hospital stroke patients).

### Intervention

The intervention used a mQIC methodology adapted from the Institute for Healthcare Improvement’s Breakthrough Series Collaborative.^19^ The mQIC began with each site formally enrolling and assembling an interdisciplinary team, as described in the *Site Enrolment and Patient Population* section. The mQIC consisted of two full-day workshops, where the enrolled team from each site attended. The first workshop highlighted evidence on treatment of ischemic stroke and strategies for improving efficiency of treatment. It included a presentation by one of the participating site’s improvement team on changes that they have implemented and are working on to improve treatment efficiency at their hospital. The afternoon consisted of each team planning their improvements and reporting it back to the entire group. The second workshop was conducted 6-8 weeks after the first, where teams presented their changes, and the afternoon also had a planning period with report back. This second workshop had presentation from four participating sites where they presented the improvements that they are working towards. Both of the workshops included a model of sharing individual site’s improvement activities, which promoted peer-to-peer learning rather than just didactive learning from experts. During the 6-month intervention period, the sites were supported with virtual site visits to each participating site. The purpose of the virtual site visits was to determine the site’s improvement teams progress towards implementing changes. Five webinars were held during the 6-month intervention, where approximately 40-60% of the webinar presentations were delivered by a participating site showing their improvement efforts. Both of the workshops and webinars had significant sharing of learning including successes and failures, activities, and action plans; additionally, they also provided presentations on evidence and best-practice. Full details of the mQIC design and engagement of all sites and personnel have previously been reported.^20^

All enrolled sites and their improvement teams were provided with a change package. This outlined the following changes for each team to trial, adapt, and implement at their site:

1. *Prenotification and stroke team activation*: paramedics pre-notify the hospital of an incoming stroke patient, and paramedics ensure two IV lines are in place; the hospital creates a single call activation of the stroke team.
2. *Rapid registration process*: the hospital adopts a “registration as *unknown*” (similar for trauma patients), pre-registration process by using information from paramedics, or a quick registration process where only minimal information is entered initially.
3. *Patient moved to CT scanner on Emergency Medical Services (EMS) stretcher*: this includes having a thrombolysis kit ready and conducting a quick neurological exam on the way to the scanner.
4. Not waiting for bloodwork when not indicated for the patient
5. *Thrombolysis administered in imaging area*
6. *Rapid transfer process for EVT*: adopting a heads-up call to the transport system when presented with a severe stroke.

### Data Collection

Data were collected by different approaches for each province. All ischemic stroke patients that received treatment with thrombolysis (alteplase and Tenecteplase) and/or EVT were included in the study for each province. NS and PE have provincial registries that contained all the necessary data. The identification of patients for inclusion (ischemic stroke patients that received acute treatment) was done in NS and PE by extracting records from their provincial stroke registries of patients that received treatment. Data is collected for the registries by a stroke coordinator through chart reviews. EMS data were linked to the registry data to obtain the date and time of the 911 call. In NB and NL, the identification of patients for inclusion was done by reviewing the dispensing of thrombolytic drug through the hospital’s pharmacy and the use of the angiosuite usage for the EVT procedure. Data from NB was collected by conducting chart reviews, which were done either by the data analyst or by the health authority’s research office. Data from NL was collected by chart review conducted either by the local stroke coordinator or through Memorial University’s Quality of Care NL organization. Although the data collection methods were heterogenous, as two provinces have a provincial registry that we were able to use for this study, data collection was done through chart reviews across all four provinces, as the provincial registries in NS and PE are populated through chart reviews. The patient chart at all participating hospitals included information on patient sex, patient age at time of stroke, onset time or last seen normal time, NIHSS, time thrombolysis was started, time that the patient left the primary stroke center for EVT, and time of arterial-access. Time of CT scan was collected through PACS (picture archiving and communication system). Data quality checks were conducted by the research team once the data was received to ensure data validity especially pertaining to the date and time fields. If there were errors discovered, they were sent back to the province for further validation. There were no exclusions made in the data collection phase. All strokes that occurred in hospital were excluded from efficiency measures: 911-to-needle, door-to-needle, 911-to-arterial-access, door-to-arterial-access, and door-in-door-out times.

### Statistical Analysis

The data for this trial includes all ischemic stroke patients in the entire province regardless of the number of sites that formally enrolled. The trial is therefore a population based study; no sample size was calculated. All missing values for age, sex, and National Institutes of Health Stroke Scale (NIHSS) were imputed using the K-Nearest Neighbor algorithm (k-NN).^25^ For the comparison analysis related to all ischemic stroke patients, unadjusted proportion comparison is conducted using χ^2^ analysis, while a mixed effects logistic regression with cluster as random effects variable is used for adjusted analysis. For analysis related to all treated patients, unadjusted analysis comparison is conducted using χ^2^ for proportions and Wilcoxon rank-sum test for continuous variables. The comparison analysis uses a mixed effects logistic regression for binary/categorical variables and a mixed effects quantile regression for continuous variables. In these analyses, age, sex, and NIHSS are fixed effects variables while cluster was a random effects variable. We did not include the time of the intervention as cluster captures the temporality of the stepped-wedge trial, and the overall intervention period is short. When analyzing results by cluster, we used a simple quantile regression model.

### Ethical Considerations and Trial Registration

Ethical approval for this study was obtained from the NS Health Research Ethics Board (REB# 1025460), Health PEI Ethics Board, Horizon Health Network Research Ethics Board (ROMEO File #: 100906 and RS#: 2020-2893), Vitalité Health Network Ethics Office (ROMEO file number: 101305), and Newfoundland and Labrador Health Research Ethics Board (Researcher Portal File #: 20210255 and Reference #: 2020.074). The trial is registered with the ISRCTN registry (ISRCTN11109800, https://www.isrctn.com/ISRCTN11109800).

## Results

Over 30 months from May 1 2020 to October 31 2022, 8594 ischemic stroke patients were included in the analysis for this trial, out of which 1576 (18.3%) patients received acute treatment. There were 3065 ischemic stroke patients in the pre-intervention period of whom 547 (17.8%) received treatment, and 3806 ischemic stroke patients were in the post-intervention period of whom 718 (18.9%) patients were treated. The full details of patient allocation by cluster and period are shown in Figures 1.

For the results of all acutely treated ischemic stroke patients, Table 1 shows the comparison of baseline characteristics. Overall the pre-intervention and post-intervention periods are similar except the median time from onset to 911 call was 5 minutes faster (p=0.001) in the post-intervention period.

**Table 1:** Baseline characteristics for all community onset ischemic stroke patients that received treatment with thrombolysis and/or endovascular thrombectomy

|  | Control |  | Intervention |  | Significance |
| --- | --- | --- | --- | --- | --- |
| **All Clusters** | **n** | **value** | **n** | **Value** |  |
| Age mean (SD) | 528 | 70.8 (13.2) | 669 | 70.4 (13.6) | 0.64 |
| Sex, women (%) | 528 | 236 (44.7%) | 670 | 303 (45.2%) | 0.90 |
| NIHSS, median (IQR) | 230 | 10 (6-14) | 242 | 10 (5-15) | 0.60 |
| Onset to door, median (IQR) | 493 | 85.0 (52.0-146.0) | 629 | 85.0 (57.0-130.0) | 0.70 |
| *Onset to 911, median (IQR)* | *364* | *22.0 (8.7-68.0)* | *434* | *17.0 (6.7-45.0)* | *0.001* |
| **Cluster 1** |  |  |  |  |  |
| Age, mean (SD) | 145 | 73.0 (13.3) | 402 | 71.01 (13.0) | 0.13 |
| Sex, women (%) | 145 | 68 (46.8%) | 402 | 188 (46.8%) | 1 |
| NIHSS, median (IQR) | 73 | 9 (5-12) | 147 | 8 (5-13) | 0.84 |
| Onset to door, median (IQR) | 144 | 84.8 (54.5-134.2) | 381 | 85.1 (60.0-127.0) | 0.94 |
| Onset to 911, median (IQR) | 99 | 14.2 (7.6-35.7) | 279 | 13.2 (6.0-35.1) | 0.40 |
| **Cluster 2** |  |  |  |  |  |
| Age, mean (SD) | 213 | 69.8 (13.7) | 179 | 68.8 (14.2) | 0.48 |
| Sex, women (%) | 213 | 95 (44.6%) | 179 | 67 (37.4%) | 0.18 |
| NIHSS, median (IQR) | 102 | 11 (7-15) | 90 | 12 (7-17) | 0.62 |
| Onset to door, median (IQR) | 192 | 82.5 (52.0-142.8) | 159 | 88.0 (55.0-135.0) | 0.71 |
| Onset to 911, median (IQR) | 167 | 25.0 (7.0-92.5) | 116 | 20.0 (7.0-63.25) | 0.36 |
| **Cluster 3** |  |  |  |  |  |
| Age, mean (SD) | 170 | 70.5 (12.2) | 89 | 70.8 (14.3) | 0.64 |
| Sex, women (%) | 170 | 73 (42.94%) | 90 | 48 (53.33%) | 0.14 |
| NIHSS, median (IQR) | 55 | 10 (6-14) | 6 | 5 (5-9.75) | 0.20 |
| Onset to door, median (IQR) | 157 | 88.0 (51.0-150.0) | 90 | 78.5 (45.0-134.0) | 0.19 |
| Onset to 911, median (IQR) | 98 | 37.5 (13.0-85.0) | 39 | 32.0 (19.0-74.0) | 0.88 |

Figure 2 shows the flow diagram for patient allocation in the pre-intervention and post-intervention periods, as well as the number of patients in the intervention period. Strokes that occurred in-hospital were excluded from the treated patient cohort, as they did not have a relevant door time thus the process measures (door-to-needle, door-to-arterial-access, and door-in-door-out time) were not relevant for these patients. Additionally, there were some community onset treated patients that had treatment times that were beyond typical standard of care, and were removed to prevent these outliers from affecting the group comparisons. These exclusions were door-to-needle time of greater than 4.5 hours; door-to-arterial-access times greater than 6 hours; and door-in-door-out times of greater than 16 hours.

**Figure 2:**
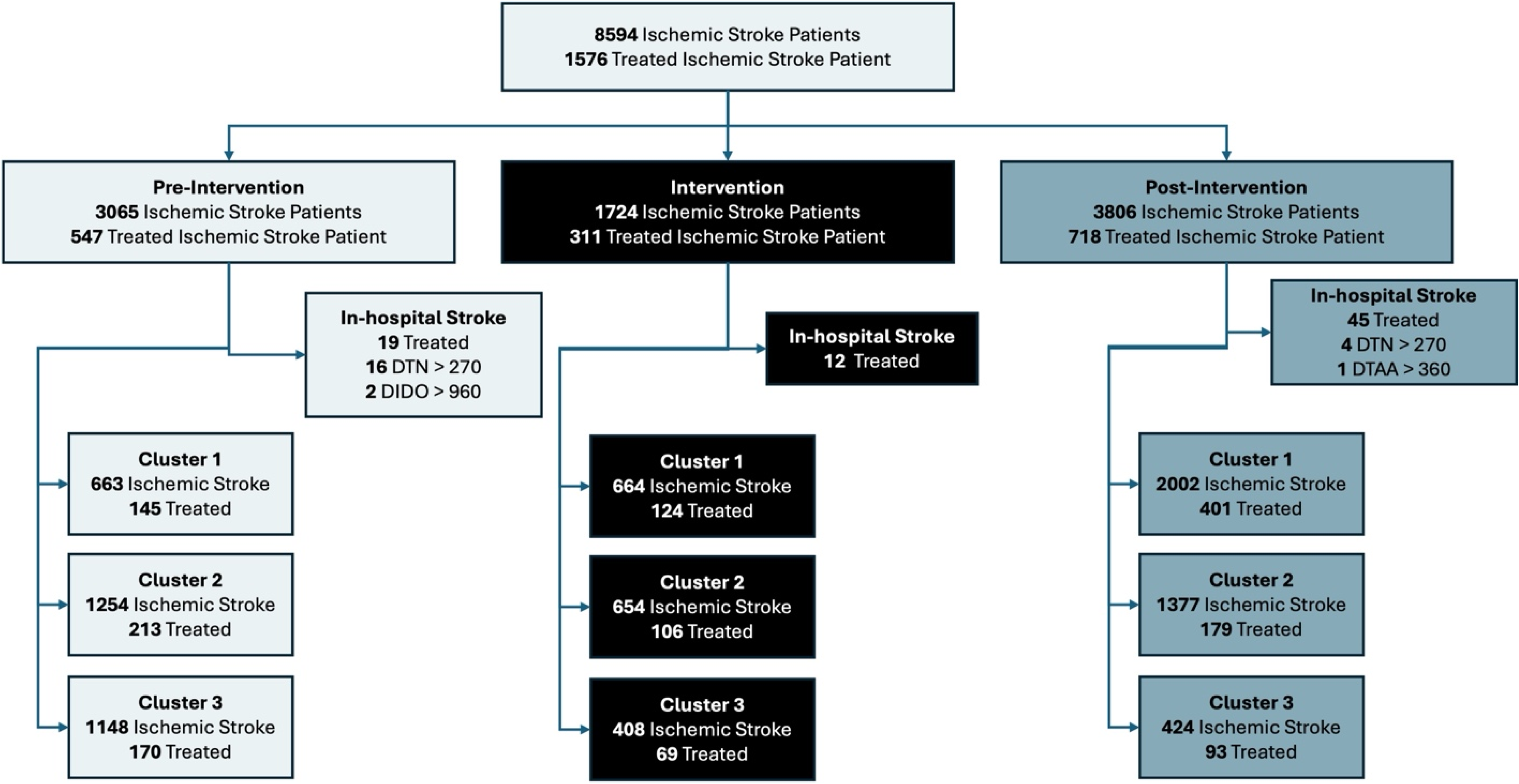
Flow diagram of patient allocation across each period and cluster. DTN: Door-to-needle time, DTAA: Door-to-arterial-access time, DIDO: Door-in-door-out time.

The results for all ischemic stroke patients are shown in Table 2. The adjusted proportion of patients that received treatment did not increase significantly with a 0.4% increase for patients that received thrombolysis and/or EVT (p=0.68). Cluster 3 showed the most significant improvements compared to the other clusters in proportion of patients treated with a 6.52% increase (p=0.003) in the proportion receiving treatment. Cluster 3 also saw a 39.35% increase (p<0.0001) in the proportion of patients discharged home. The trends for the proportion of ischemic stroke patients that received treatment is shown in Figure 3. Cluster 1 and 3 showed upward trends while cluster 2 showed a downward trend.

**Figure 3:**
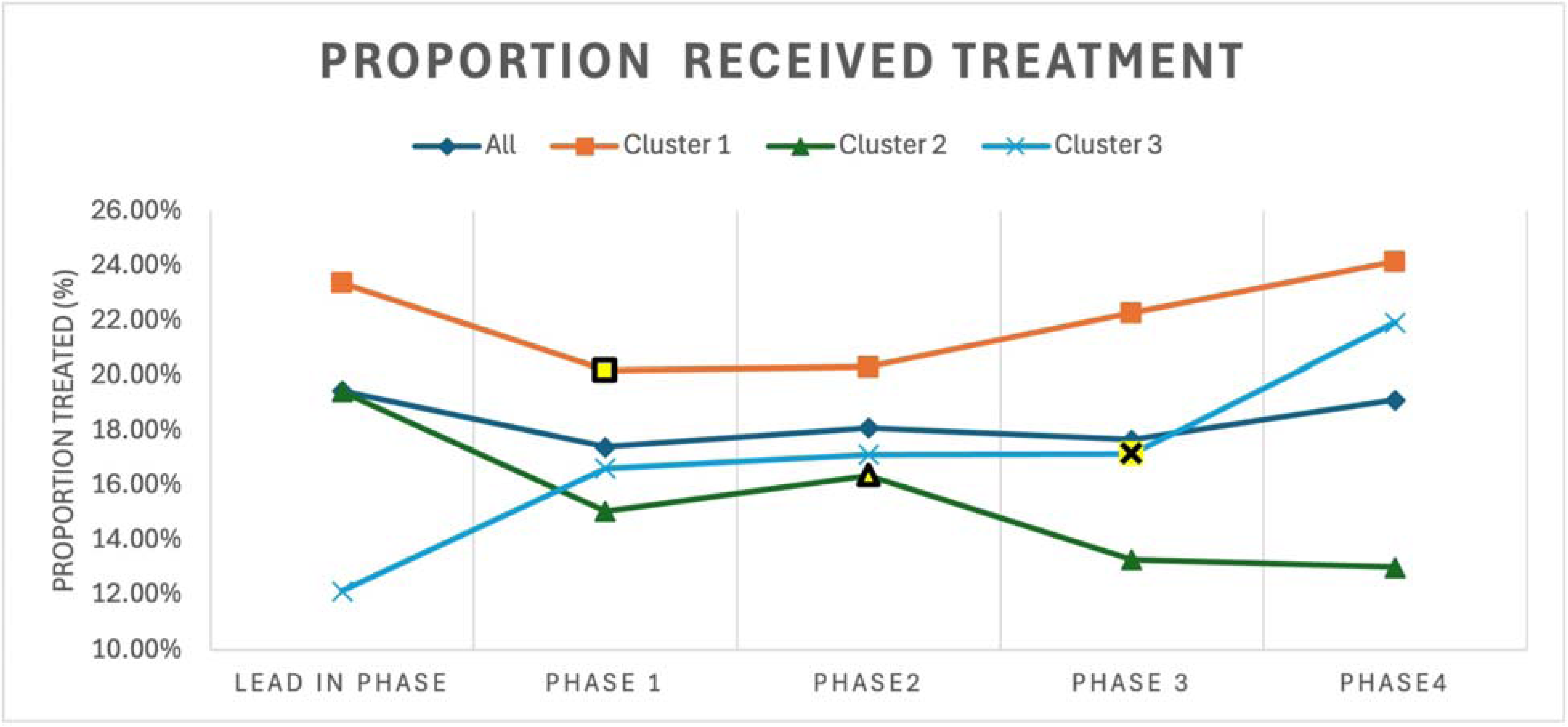
Trends for proportion of ischemic stroke patients that received treatment for the entire cohort and for each cluster by trial phase. The intervention period for each cluster is highlighted in yellow.

**Table 2:** Proportion of patients treated and discharge disposition for all ischemic stroke patients; adjustments are only made for cluster allocation; treated patients include both community and hospital onset stroke patients.

| Entire Cohort | Pre period (Control)<br>n = 3065 | Post period<br>(Intervention)<br>n = 3803 | p (significance) |
| --- | --- | --- | --- |
|  | N (%) | N (%) |  |
| Received Thrombolysis | 474 (15.46%) | 620 (16.30%) | 0.36 |
| Received Thrombolysis (adjusted) | 495 (16.15%) | 647 (17.02%) | 0.3 |
| Received EVT | 115 (3.75%) | 196 (5.15%) | 0.006 |
| Received EVT (adjusted) | 123 (4.00%) | 151 (3.98%) | 0.9 |
| Received either thrombolysis and/or EVT | 547 (17.85%) | 718 (18.87%) | 0.28 |
| Received either thrombolysis and/or EVT (Adjusted) | 570 (18.60%) | 723 (19.00%) | 0.68 |
| Transferred for EVT | 118 (3.84%) | 193 (5.07%) | 0.017 |
| Transferred for EVT (Adjusted) | 127 (4.15%) | 145 (3.80%) | 0.14 |
| Discharged home | 1015 (33.10%) | 1569 (41.25%) | < 0.0001 |
| *Discharged home (Adjusted)* | *1016 (33.15%)* | *1768 (46.50%)* | *< 0.0001* |
| Died | 324 (10.57%) | 570 (14.98%) | < 0.0001 |
| Died (Adjusted) | 349 (11.39%) | 464 (12.20%) | 0.3 |
| **Cluster 1** | **N=663** | **N=2002** |  |
| Received Thrombolysis | 132 (19.90%) | 383 (19.10%) | 0.70 |
| Received EVT | 40 (6.03%) | 122 (6.09%) | 1 |
| Received either thrombolysis and/or EVT | 155 (23.37%) | 444 (22.17%) | 0.55 |
| Transferred for EVT<sup>®</sup> | 43 (6.40%) | 114 (5.60%) | 0.51 |
| Discharged home | 236 (35.59%) | 699 (34.91%) | 0.78 |
| Died | 117 (17.64%) | 360 (17.98%) | 0.89 |
| **Cluster 2** | **N=1254** | **N=1377** |  |
| Received Thrombolysis | 165 (13.15%) | 147 (10.67%) | 0.056 |
| Received EVT | 75 (5.98%) | 71 (5.15%) | 0.40 |
| Received either thrombolysis and/or EVT | 215 (17.14%) | 181 (13.14%) | 0.004 |
| Transferred for EVT<sup>®</sup> | 75 (5.98%) | 79 (5.73%) | 0.85 |
| Discharged home | 543 (43.30%) | 616 (44.73%) | 0.48 |
| Died | 165 (13.15%) | 189 (13.72%) | 0.71 |
| **Cluster 3** | **N=1148** | **N=424** |  |
| *Received Thrombolysis* | *177 (15.41%)* | *90 (21.22%)* | *0.0081* |
| *Received EVT* | *0* | *3 (0.70%)* | *0.02* |
| *Received either thrombolysis and/or EVT* | *177 (15.41%)* | *93 (21.93%)* | *0.003* |
| Transferred for EVT | 0 | 0 | - |
| *Discharged home* | *236 (20.55%)* | *254 (59.90%)* | *< 0.0001* |
| Died | 42 (3.65%) | 21 (4.95%) | 0.30 |

The process and outcomes results for all community onset ischemic patients treated with thrombolysis and/or EVT are shown in Table 3. The only time measure to see a significant reduction is door-to-needle time, which went down to a median of 62.0 minutes (IQR: 38.7-85.9 minutes) from 71.2 minutes (IQR: 43.5-99.5 minutes) (p=0.01). Cluster 1 showed no significant improvements. Cluster 2 showed a 7.5 minute reduction in the median door-to-CT time (p=0.009), and a 12.6 minute reduction in the median door-to-needle time (p=0.01). Cluster 3 showed the most improvement, and their post-intervention median door-to-needle times was the lowest of all 3 clusters; they went from a median door-to-needle time of 54.5 minutes (IQR: 44.0-67.0 minutes) to a median of 49.0 minutes (IQR: 41.7-56.3 minutes) (p=0.05). This improvement also carried through to the median 911-to-needle time, where the median time dropped by 12.7 minutes (p=0.05). Cluster 3 also saw a 5.81% increase (p=0.0081) in the proportion of ischemic stroke patients that received thrombolysis, and a 6.52% increase (p=0.003) in the proportion of ischemic stroke patients that received thrombolysis and/or EVT. The trends for all acutely treated ischemic stroke patients are shown in Figure 4. Panel A show the trends for median door to needle time for all thrombolysed patients with cluster 2 and 3 showing a downward trend. Panel B shows the trend for median door-in-door-out times with phase 2 showing longer times across all clusters. Panel C shows the trend for median door-to-arterial-access times with cluster 1 showing a downward trend. Note that the trends for door-in-door-out and door-to-arterial-access times do not include cluster 3, as they only started their EVT service in the final phase. Panel D on Figure 4 show the trends for proportion of treated ischemic stroke patients that are discharged home.

**Figure 4:**
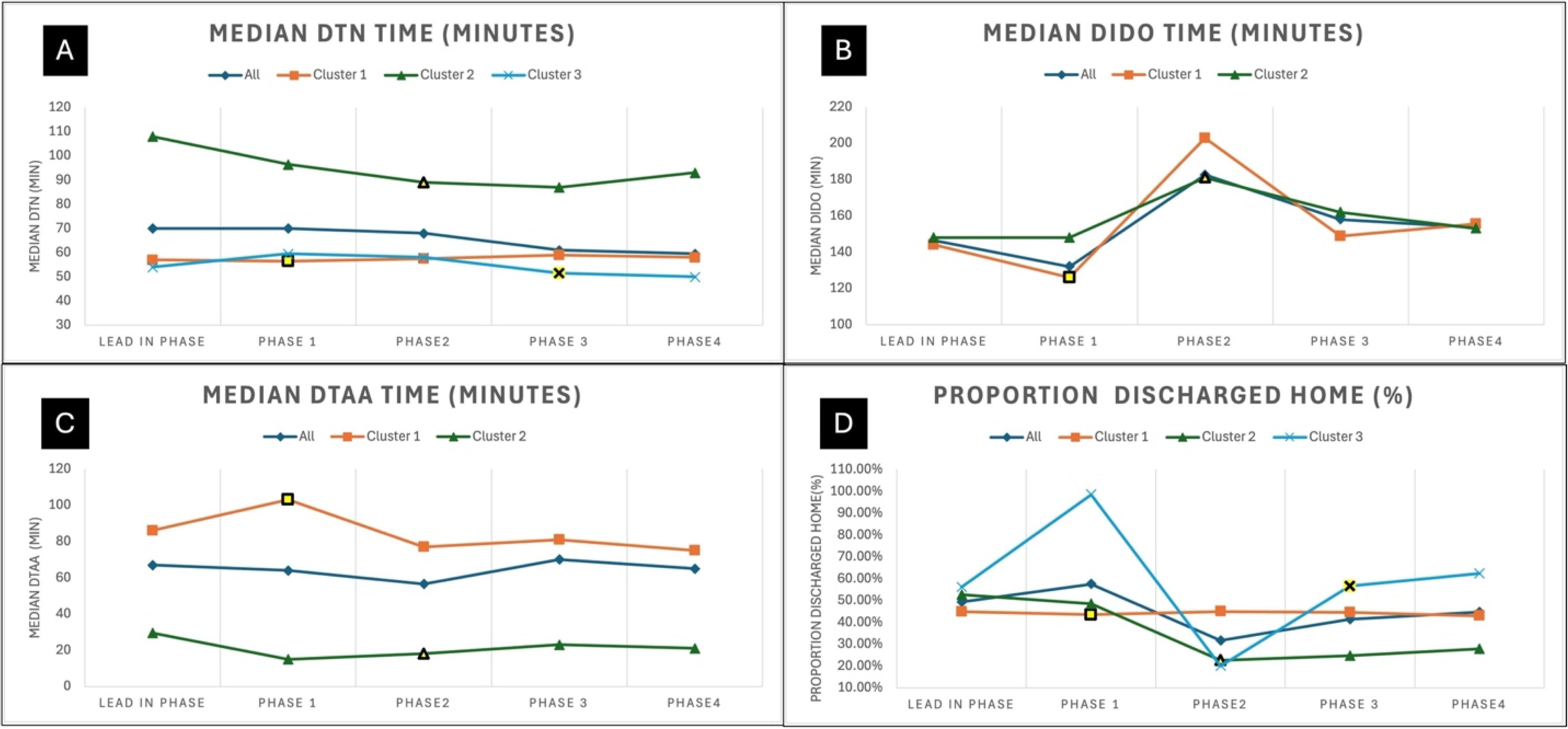
Trends for community onset ischemic stroke patients that received treatment for each phase. Panel A shows the trends for median door-to-needle time for all thrombolysed ischemic stroke patients. Panel B shows the trends for median door-in-door-out time for all patients transferred for EVT. Panel C shows the trends for median door to Arterial Access time for all patients treated with EVT. Panel D shows the trends for proportion of community onset treated ischemic stroke patients discharged home from acute care. The intervention period for each cluster is highlighted in yellow. EVT: Endovascular thrombectomy, DTN: Door-to-needle time, DTAA: Door-to-arterial-access time, DIDO: Door-in-door-out time.

**Table 3:**
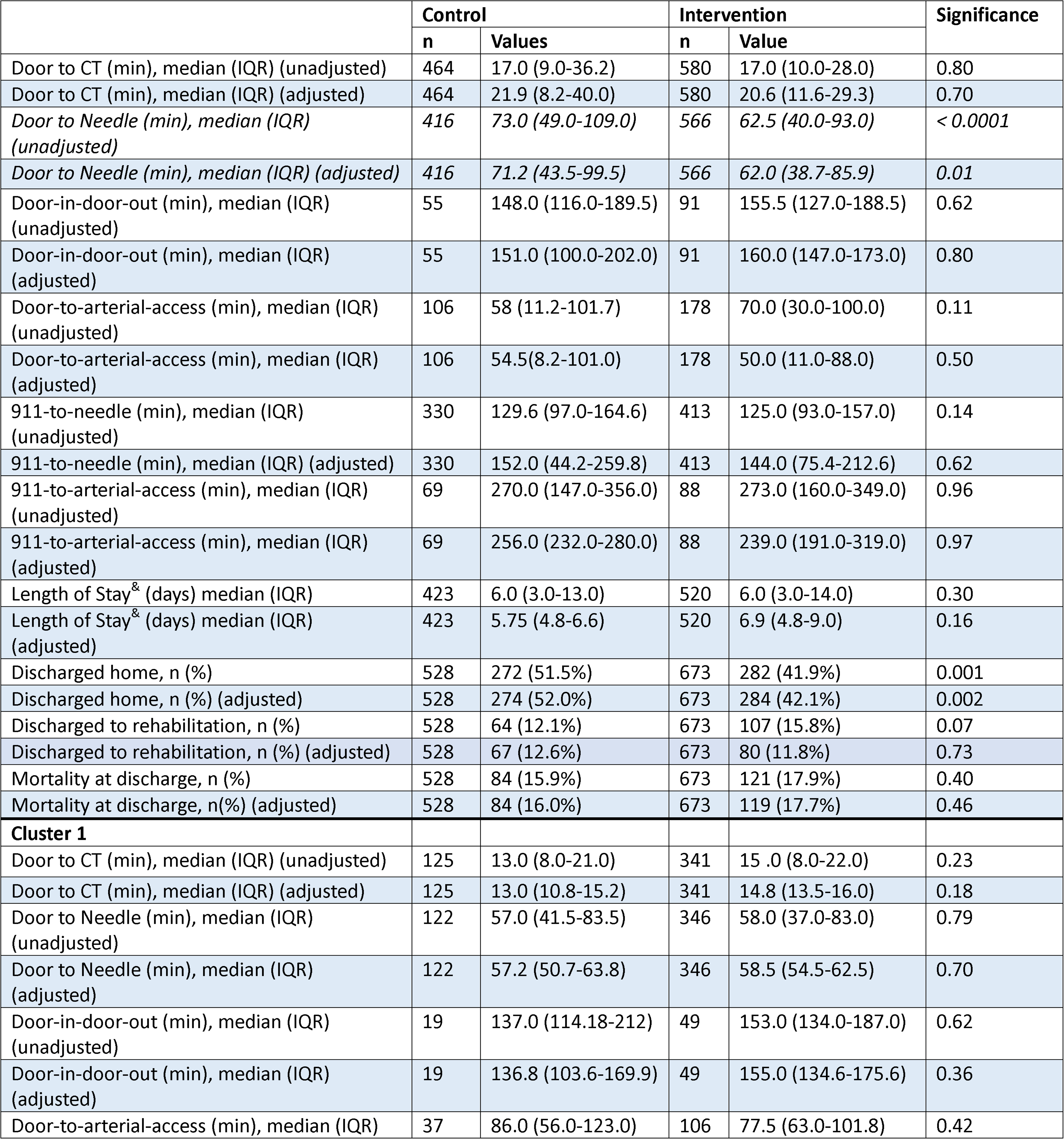
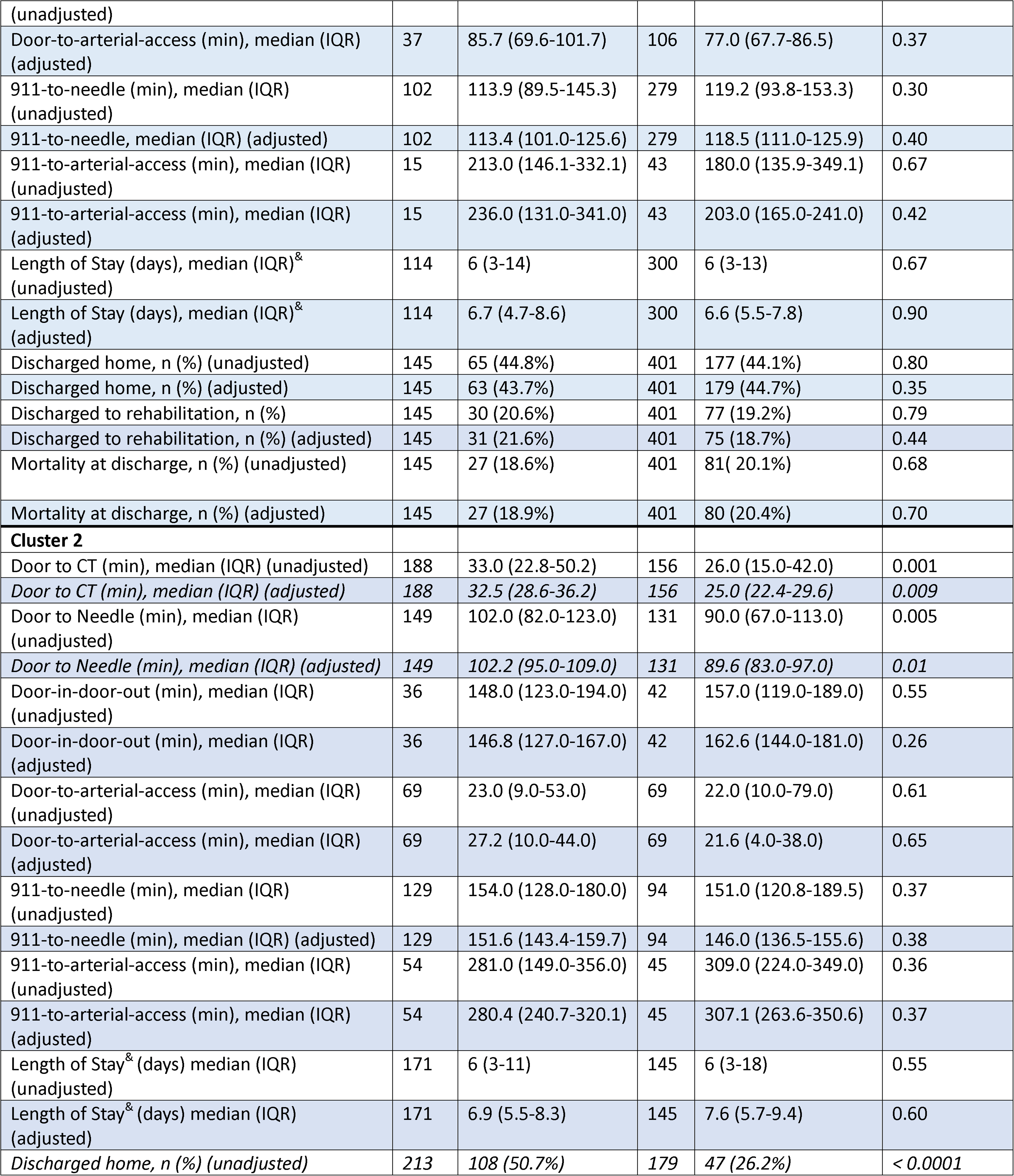
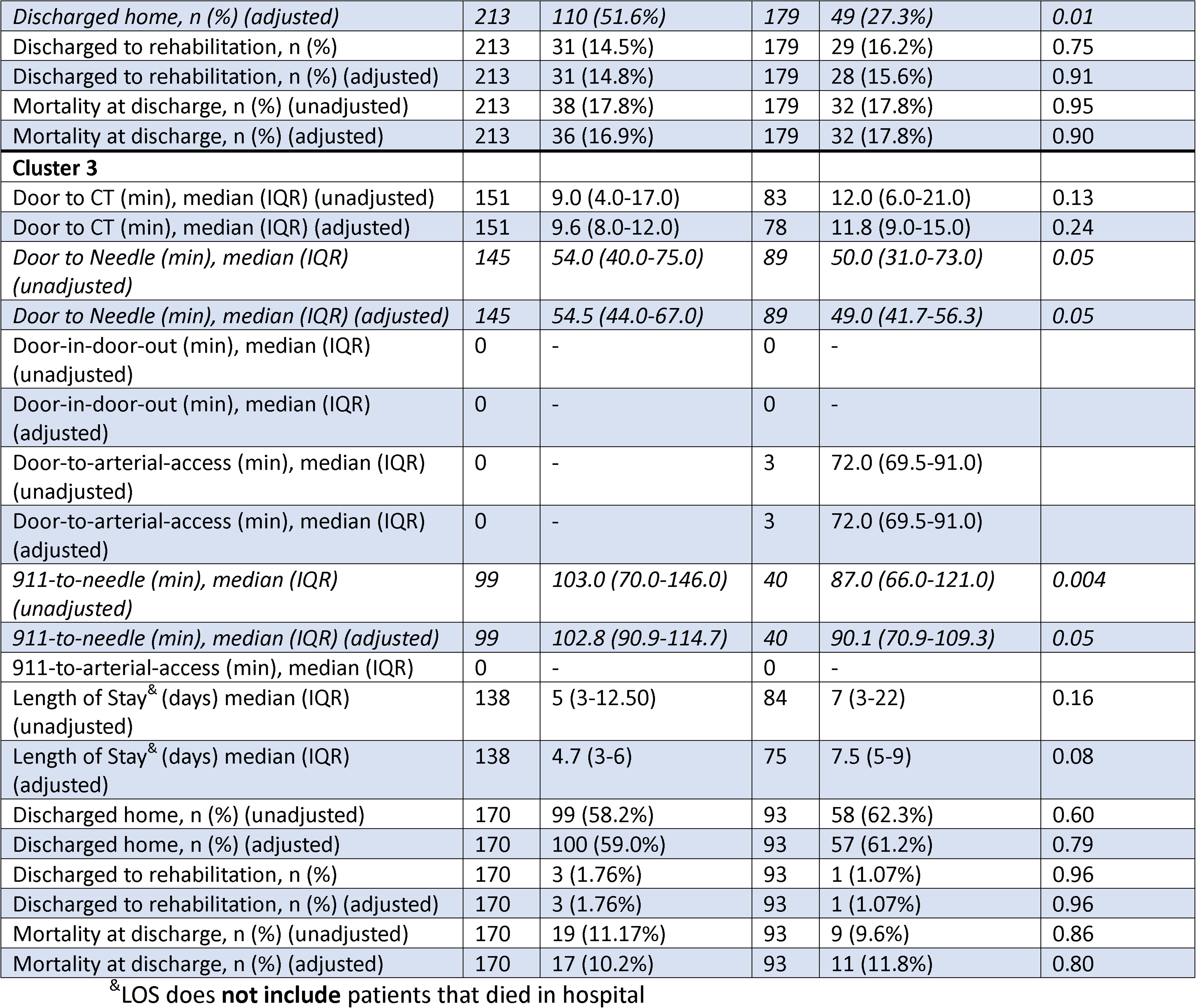
Treatment times and outcome measures for all community onset ischemic stroke patients treated with thrombolysis and/or endovascular thrombectomy. Adjustments were made for cluster, age, sex and NIHSS. IQR: interquartile range, min: minutes.

## Discussion

These results show that improvements were minimal and highly dependent on the cluster. Although there were improvements to the door-to-needle times, these improvements are only present in clusters 2 and 3. However, the trends reveals additional details, as Figure 3 shows that cluster 1 and 2 showed an increasing trends in the proportion of ischemic stroke patients that received treatment after the intervention, which suggests that the intervention had an positive effect in improving proportion treated for these two clusters; however, cluster 2 showed a downward trend, which resulted in no overall affect across the clusters. It is difficult to determine why cluster 2 showed this negative affect in the proportion treated, as this cluster also has the lowest proportion of ischemic stroke patients treated across the clusters.

The results for the efficiency of treatments showed the overall reduction in median door-to-needle time was 9.2 minutes. The trends show that cluster 2 and 3 trended downwards. However, cluster 2 is showing that the last period did trend upwards, so the improvements may not be sustained. The door-in-door-out time times showed no difference across clusters or impact of the intervention on this measure. Additionally, phase 2 show a stark increase in door-in-door-out times across the two clusters that were transferring patients, which may have been due to increased COVID-19 measures in the region at that time. Interestingly, cluster 1 showed a downward trend after the intervention for door-to-arterial-access time, which appears to have been sustained, which was not statistically significant, but a potential positive impact of the intervention.

The overall reduction in door-to-needle time of 9.2 minutes is much less compared to previous studies that used a similar intervention.^26–28^ The door-to-needle initiative in Alberta saw approximately 30-minute reduction in median times;^26^ the California initiative resulted in approximately 20-minute reduction;^27^ and the Chicago initiative saw a 15.5 minute reduction.^28^ None of these studies used a stepped-wedge trial design, which may account for some of the lower impact, as their improvements may have been overstated due to secular trends and other factors. The settings in Alberta, California, and Chicago studies are much more urban than Atlantic Canada; even in the population study for Alberta Canada, where 17% of their population lives in rural areas compared to approximately 50% for Atlantic Canada.^21^ Additionally, the intervention period was much shorter in this study, which may suggest that more time is needed to allow sites to implement their changes especially if more patients are being seen outside of urban centers. Furthermore, the entire trial took place during a time of COVID-19 restrictions. Hospitals had additional tasks such as conducting a COVID-19 test on all patients; furthermore, personnel were participating in projects related to mitigating spread of COVID-19, which created a significant distraction from implementing changes to improve acute stroke treatment.^29^ In addition, the mQIC was held entirely virtually due to the pandemic, and these results may signal that virtual Improvement Collaboratives are less efficacious, and there is a need for face-to-face workshops and site visits. Previous studies have noted that virtual quality improvement require a longer lead time,^30^ so a shorter implementation period combined with virtual delivery in our study may have had a compounding effect on the impact of the intervention.

A significant limitation of the intervention was that key process data was not available during the intervention period. Data was collected for the purpose of this trial after the intervention period due to the significant effort to extract the data from the charts. Even the provinces with stroke registries had significant delays in the recording of the data. This resulted in improvement efforts being conducted without data feedback, which is critical for improvement. This may have been another reason why there was lower impact of the intervention from previous studies that used a similar intervention, as one of the previous studies developed a provincial registry for quality improvement efforts during their QIC.^26^

These results show that the intervention also needs to be tailored for each health system. This study shows that cluster 3 was able to improve the proportion of patients that receive treatment and efficiency of treatment, cluster 2 was able to improve door-to-needle times, and cluster 1 was not able to make any changes. Since this stepped-wedge trial was designed such that each intervention was similar with limited customization, this may have had variable impact on each health system. This shows the need to assess each health system and tailor the implementation strategy for each health system based on their barriers.^31^ Furthermore, additional customized supports to each site may have been necessary, which was not done in this intervention.

The process changes were observed in the door-to-needle metric with the trends showing a positive impact for door-to-arterial-access for cluster 1. This is likely because the improvement strategies were based on improving this single metric. Additional evidence-based strategies to improve door-to-arterial-access and door-in-door-out times are needed to see improvements in these metrics. Furthermore, Improvement Collaboratives that focus on a single metric are shown to be more successful than those that focus on multiple metrics and improvements.^26, 32^

To summarize, the mQIC intervention should be enhanced to observe greater improvements. The suggested enhancements are: 1) the intervention period should be a minimum of 1-year long to allow teams enough time to make the changes; 2) the workshops and site visits should be in-person rather than virtual; 3) data feedback needs to occur throughout the intervention; 4) pre-intervention readiness assessments should be completed for each cluster to tailor the intervention for each cluster; and 5) change suggestions should include ways to lower door-in-door-out times and door-to-groin-puncture times.

Although the results showed significant improvements to the proportion of patients discharged home, it is unlikely that the intervention had this level of impact on this pragmatic outcome measures. Although there was an increase in the proportion of patients that received treatment in cluster 3 and improvement in thrombolysis time for clusters 2 and 3, these improvements are unlikely to have seen a 13% improvement in proportion of ischemic stroke patients discharged home. This shows the need to have true outcomes measures such as 90-day mRS (modified Rankin Score) even for quality improvement initiatives.

## Conclusions

The intervention to improve access and efficiency of acute ischemic stroke treatment in four Atlantic Canadian provinces made a 9.2-minute improvement in median door-to-needle times. A longer intervention period that is 1-year long and tailored to each health system with in-person workshops and sites visits may result in more improvements.

## Supporting information

CONSORT Checklist

Protocols

## Data Availability

All data were obtain through data sharing agreements between the health system and Dalhousie University, and the data cannot be shared outside of this agreement.

## Acknowledgements

Owen Parfrey at Memorial University’s Quality of Care NL assisted with facilitating data procurement and collection across NL. Jody Rhodenizer at Horizon Health Network was key in obtaining data for all Horizon hospital in NB. We thank the research office at Vitalité Health Network for visiting hospitals in the Vitalité Health Authority in NB. Dr. Jessalyn Holodinsky guided us in understanding the best methodology for statistical analysis.

