## Supplementary material for "Improving Access and Efficiency of Acute Ischemic Stroke Treatment Across Four Canadian Provinces: A Stepped-Wedge Trial": CONSORT Checklist

**Supplementary materials 3: Checklist of information to include when reporting a stepped wedge cluster randomised trial (SW-CRT)**

| Topic | Item no | Checklist item | Page no |
|---|---|---|---|
| **Title and abstract** | | | |
| | 1a | Identification as a SW-CRT in the title. | 1 |
| | 1b | Structured summary of trial design, methods, results, and conclusions (see separate SW-CRT checklist for abstracts). | 1,4 |
| **Introduction** | | | |
| Background and objectives | 2a | Scientific background. Rationale for using a cluster design and rationale for using a stepped wedge design. | 5,6 |
| | 2b | Specific objectives or hypotheses. | 6 |
| **Methods** | | | |
| Trial design | 3a | Description and diagram of trial design including definition of cluster, number of sequences, number of clusters randomised to each sequence, number of periods, duration of time between each step, and whether the participants assessed in different periods are the same people, different people, or a mixture. | 6,20 |
| | 3b | Important changes to methods after trial commencement (such as eligibility criteria), with reasons. | N/A |
| Participants | 4a | Eligibility criteria for clusters and participants. | 8,11 |
| | 4b | Settings and locations where the data were collected. | 7 |
| Interventions | 5 | The intervention and control conditions with sufficient details to allow replication, including whether the intervention was maintained or repeated, and whether it was delivered at the cluster level, the individual participant level, or both. | 9-10 |
| Outcomes | 6a | Completely defined prespecified primary and secondary outcome measures, including how and when they were assessed. | 8-9 |
| | 6b | Any changes to trial outcomes after the trial commenced, with reasons. | N/A |
| Sample size | 7a | How sample size was determined. Method of calculation and relevant parameters with sufficient detail so the calculation can be replicated. Assumptions made about correlations between outcomes of participants from the same cluster. (see separate checklist for SW-CRT sample size items). | 11 |
| | 7b | When applicable, explanation of any interim analyses and stopping guidelines. | N/A |
| **Randomisation** | | | |
| Sequence generation | 8a | Method used to generate the random allocation to the sequences of treatments. | 6 |
| | 8b | Type of randomisation; details of any constrained randomisation or stratification, if used. | N/A |
| Allocation concealment mechanism | 9 | Specification that allocation was based on clusters; description of any methods used to conceal the allocation from the clusters until after recruitment. | 6 |
| Implementation | 10a | Who generated the randomisation schedule, who enrolled clusters, and who assigned clusters to sequences. | 6 |
| | 10b | Mechanism by which individual participants were included in clusters for the purposes of the trial (such as complete enumeration, random sampling; continuous recruitment or ascertainment; or recruitment at a fixed point in time), including who recruited or identified participants. | 6,11 |
| | 10c | Whether, from whom and when consent was sought and for what; whether this differed between treatment conditions. | N/A |
| Blinding | 11a | If done, who was blinded after assignment to sequences (eg, cluster level participants, individual level participants, those assessing outcomes) and how. | N/A |
| | 11b | If relevant, description of the similarity of treatments. | N/A |
| Statistical methods | 12a | Statistical methods used to compare treatment conditions for primary and secondary outcomes including how time effects, clustering and repeated measures were taken into account. | 11 |
| | 12b | Methods for additional analyses, such as subgroup analyses, sensitivity analyses, and adjusted analyses. | 11 |

(*Continued*)

**Supplementary materials 3 (*Continued*)**

| Topic | Item no | Checklist item | Page no |
|---|---|---|---|
| **Results** | | | |
| Participant flow (a diagram is strongly recommended) | 13a | For each treatment condition or allocated sequence, the numbers of clusters and participants who were assessed for eligibility, were randomly assigned, received intended treatments, and were analysed for the primary outcome (see separate SW-CRT flow chart). | 21 |
| | 13b | For each treatment condition or allocated sequence, losses and exclusions for both clusters and participants with reasons. | 21 |
| Recruitment | 14a | Dates defining the steps, initiation of intervention, and deviations from planned dates. Dates defining recruitment and follow-up for participants. | 20 |
| | 14b | Why the trial ended or was stopped. | N/A |
| Baseline data | 15 | Baseline characteristics for the individual and cluster levels as applicable for each treatment condition or allocated sequence. | 12,23 |
| Numbers analysed | 16 | The number of observations and clusters included in each analysis for each treatment condition and whether the analysis was according to the allocated schedule. | 12,20,21 |
| Outcomes and estimation | 17a | For each primary and secondary outcome, results for each treatment condition, and the estimated effect size and its precision (such as 95% confidence interval); any correlations (or covariances) and time effects estimated in the analysis. | 12,13,22 |
| | 17b | For binary outcomes, presentation of both absolute and relative effect sizes is recommended. | N/A |
| Ancillary analyses | 18 | Results of any other analyses performed, including subgroup analyses and adjusted analyses, distinguishing prespecified from exploratory. | N/A |
| Harms | 19 | Important harms or unintended effects in each treatment condition (for specific guidance see CONSORT for harms). | N/A |
| **Discussion** | | | |
| Limitations | 20 | Trial limitations, addressing sources of potential bias, imprecision, and, if relevant, multiplicity of analyses. | 13-14 |
| Generalisability | 21 | Generalisability (external validity, applicability) of the trial findings. Generalisability to clusters or individual participants, or both (as relevant). | 14 |
| Interpretation | 22 | Interpretation consistent with results, balancing benefits and harms, and considering other relevant evidence. | 14 |
| **Other information** | | | |
| Registration | 23 | Registration number and name of trial registry. | 11 |
| Protocol | 24 | Where the full trial protocol can be accessed, if available. | 11 |
| Funding | 25 | Sources of funding and other support (such as supply of drugs), and the role of funders. | 15-16 |
| Research ethics review | 26 | Whether the study was approved by a research ethics committee, with identification of the review committee(s). Justification for any waiver or modification of informed consent requirements. | 11 |

This checklist has been taken from table 3 in *BMJ* 2018;363:k1614, as a standalone document for readers to print out or fill in electronically.
