## Supplementary material for "Improving Access and Efficiency of Acute Ischemic Stroke Treatment Across Four Canadian Provinces: A Stepped-Wedge Trial": Protocols

### Research Protocol

**STUDY TITLE:** **Atlantic Canada Together Enhancing Acute Stroke Treatment (ACTEAST): Improving Access and Efficiency of Treatment**

**CLINICAL STUDY REGISTRATION NUMBER:** *ISRCTN11109800*

**PRINCIPAL INVESTIGATOR:**

***Noreen Kamal***
Assistant Professor
Department of Industrial Engineering
Dalhousie University
5269 Morris Street, Room 100
PO Box 15000
Halifax NS  B3H 4R2 Canada
+1 (902) 494-3293


**CO-INVESTIGATORS:**

Stephen Phillips, NSHA/Dalhousie University,
NS Lead
Gordon Gubitz, NSHA/Dalhousie University,
Alier Marrero, Vitalité Health Network,
NB (Vitalité Health Network) Lead
Tania Chandler, Horizon Health Network,
NB Lead
Brian Archer, Horizon Health Network,
Michael Hill, University of Calgary,
Bijoy Menon, University of Calgary,
Kelly Mrklas, Alberta Health Services,
Peter Vanberkel, Dalhousie University,
John Blake, Dalhousie University,
Greg Browne, Memorial University,
Etienne Van Der Linde, Eastern Health,
Emergency Medicine Lead
Brian Metcalf, Eastern Health,
Fraser Clift, Memorial University,
NL Lead
Alix Carter, Dalhousie University,
Judah Goldstein, Dalhousie University,
Patrick Fok, Dalhousie University,
David Volders, NSHA/Dalhousie University,
Jens Heidenreich, NSHA/Dalhousie University,
Health Williams, PEI Health,
PEI Lead
Adela Cora, NSHA/Dalhousie University,
Dylan Blacquiere, University of Ottawa,

**STUDY SPONSOR:** none

**FUNDER:** This study is being funded by CIHR Project Grant

**Background on Stroke Treatment:**
Revolutionary advances in stroke treatment have occurred over the past two decades from thrombolysis using IV (intravenous) alteplase in the mid-1990's[1,2] to the most recent ground-breaking trials showing the high efficacy of endovascular thrombectomy (EVT) in 2015.[3,4] Studies demonstrate that an additional 10% of patients receiving alteplase within 3 hours of stroke onset will have no disability compared to control, which translates to a number needed to treat (NNT) of 10.[5] EVT is even more efficacious, with studies showing that an additional 14% of patients will have no disability and 20% will be functionally independent (NNT=5).[3] These two synergistic therapies are used to treat acute ischemic stroke, which is the most common type of stroke, making up 85% of all strokes. EVT is provided to a subset of ischemic stroke patients with the most severe form of ischemic stroke due to a large vessel occlusion, while alteplase is appropriate for a larger proportion of ischemic stroke patients including those with both large and small vessel occlusions.

Stroke is the leading cause of severe disability,[6] which has significant societal and economic impact. Stroke results in disability that increases the need for assistance with daily living tasks,[7] impedes return to the workplace,[8] and results in high hospital costs. These high hospital costs are related to extended hospitalization during the acute phase, prolonged in-patient rehabilitation, and the need for long-term care.[9] Therefore, because the therapies attenuate the severity of disability, the therapies reduce costs and provide significant societal benefit.

*Evidence-to-Practice Gap:* Although alteplase and EVT are part of guideline care in Canada and around the world,[10,11] less than optimal utilization rates for both of these treatments are observed. This evidence-to-practice gap is not a new phenomenon;[12,13] however, the substantial economic and societal benefits of these therapies make it critical to pursue optimal uptake in Atlantic Canada and the rest of the country.

This evidence-practice gap is exacerbated by the geo-political and socio-political divide. There is a split between urban and rural access to treatment. Emergency physicians are less comfortable with intravenous alteplase,[14-16] and they are often the only treating physician in rural hospitals, where access to stroke physicians or neurologists is limited. In fact, the Canadian Association of Emergency Physician only endorsed alteplase in 2015 (revised in 2018) and only to 3 hours after onset.[17] Atlantic Canada is especially challenged by access gaps. The Queen Elizabeth II Health Sciences Centre (QEII) provides guideline- and evidence-based treatment to the residents of metropolitan Halifax, where 22.8% of ischemic stroke patients received alteplase in 2018, but in rural Nova Scotia only 17.5% received alteplase. In New Brunswick, similar discrepancies exist with 15.7% receiving alteplase in Saint John and Moncton, and only 4.8% receiving alteplase outside of these cities (2017/18). In PEI (Prince Edward Island), 9% received alteplase in 2017. The alteplase rates in Newfoundland and Labrador (NL) were 8.7% in 2017/18.

Access to EVT for rural areas is even more challenging because the treatment is new and requires specialised equipment and personnel. This is particularly true in Atlantic Canada where EVT is only provided in Halifax, NS (QEII), and Saint John, NB. In metropolitan Halifax, 9% of ischemic stroke patients received EVT, but in rural Nova Scotia only 1% received EVT (2017). PEI transfers patients to Halifax and, in some cases, to Saint John, but only 2% of ischemic stroke patients were transferred and only 1% received EVT upon arrival. EVT is not yet available in NL.

Although both of these therapies mitigate disability, their effectiveness has been shown to be highly time dependent. In stroke treatment, minutes matter,[5,18] bringing to light the mantra, "time is brain".[19] Therefore, there have been calls to reduce hospital alteplase treatment times to 30 minutes from hospital arrival,[10,20] and to create seamless transfer processes for more efficient access to EVT for patients living outside major urban centres.[21,22] In Atlantic Canada, alteplase treatment timeliness does not meet the benchmark of 30 minutes (median time to treatment, Door-to-Needle time, DNT). The DNT in Nova Scotia is 67 minutes and 92 minutes in PEI. In New Brunswick, the median DNT is 100 minutes. DNT in NL is not available.

*The Context:* Stroke treatment in Atlantic Canada is particularly challenging because small populations are dispersed over wide rural areas. Atlantic Canada has a much larger percent of its population that live in rural areas: in Nova Scotia 43% of its population live in rural areas, 48% in NB; 53% in PEI; and 41% in NL. This is more than double other Canadian provinces; for example, only 14%, 17%, 14%, and 19% live in rural areas for Ontario, Alberta, British Columbia, and Quebec, respectively.[23] Atlantic Canada also has an older population.[24] The age and risk standardized stroke incidence per 100,000 people is greater in Atlantic Canada than in other provinces: up to 140 strokes/100,000 people in the Atlantic provinces compared to 113 strokes/100,000 people in Ontario.[25]

Contributing to the geographic and rural challenges is provincial responsibility for delivery of health care, which impedes the implementation of evidence-based practice trans-provincially. The creation of interprovincial protocols is challenging because health care delivery and budgets are managed provincially with no incentive to create cross-provincial systems, even though the outcomes of such programs would benefit the entire population of the region. A concerted effort to develop patient-focused processes is necessary.

**Objectives, Hypotheses and Measures:**
*The ACTEAST (Atlantic Canada Together Enhancing Acute Stroke Treatment) project aims to improve access and efficiency of stroke treatment across Atlantic Canada*. Specifically, ACTEAST will study the effectiveness of the proposed intervention through both a rigorous quantitative study using a quasi-experimental stepped-wedge design and qualitative inquiry. There is an imperative to study the proposed quality improvement intervention in a Canadian context within the Atlantic provinces because of its local political and geographic attributes. The primary hypothesis for the ACTEAST project is the following:

*The intervention will* ***increase the proportion of ischemic stroke patients that receive either alteplase or EVT by 5%.***

The secondary hypotheses are as follows:

- The components of the intervention that were most effective in facilitating improvements will be determined (qualitative)
- The intervention will reduce the median DNT of all alteplase treated patients
- The intervention will increase the proportion of all ischemic stroke patients that are discharged home from acute care
- The intervention will increase the proportion of treated ischemic stroke patients that are discharged home from acute care
- The intervention will reduce the hospital length of stay for all ischemic stroke patients
- The intervention will reduce the hospital length of stay for treated ischemic stroke patients
- The intervention will reduce the door-in-door-out times for all patients transferred for EVT
- The intervention will reduce the door-to-groin-puncture times for all EVT treated patients

- The intervention will reduce time to treatment from first medical contact (911 call)
- The Learning Sessions were effective (using the evaluations from all Learning Sessions)

*Objectives:* The overarching research objective is to enhance both access to, and efficiency of, stroke treatment in order to improve outcomes for patients across Atlantic Canada. The primary objective is to:

- Increase the percent of ischemic stroke patients that receive either alteplase or EVT by 5%

The key secondary objective is to:

- Reduce the DNT times to a median of 30 minutes, and increase the percent of patients that receive alteplase within 60 minutes to 90%[19]

These "stretch" targets have the potential to create substantive improvements beyond what "realistic" targets would achieve, based on the experience of a similar intervention in Alberta that was led by N. Kamal. The measures for these objectives are shown in the table below:

| Hierarchy | Measure | Goal | Source measures |
|---|---|---|---|
| **Primary** | Proportion of ischemic stroke patients receiving alteplase | Increase percent of ischemic stroke patients that receive treatment by 5% | - # of patients treated with alteplase<br>- # of ischemic stroke patients |
| **Primary** | Proportion of ischemic stroke patients receiving EVT | | - # of patients treated with EVT<br>- # of ischemic stroke patients |
| **Primary** | Door-to-Needle Time (DNT) | Median Door-to-Needle time under 30 minutes; 90% within 60 minutes | - time of alteplase start<br>- time of arrival |
| **Secondary** | Proportion of all ischemic stroke patients discharged home (increase) | | - # of ischemic stroke patients discharged home from acute care<br>- # of ischemic stroke patients |
| **Secondary** | Proportion of treated (alteplase or EVT) ischemic stroke patients discharged home (increase) [adjusted for age, sex and stroke severity to account for imbalance between groups] | | - # of treated ischemic stroke patients discharged home from acute care<br>- # of treated ischemic stroke patients |
| **Secondary** | Hospital length-of-stay for all ischemic stroke patients (reduce) | | - date of discharge from hospital (including in-patient rehabilitation)<br>- date of admission |
| **Secondary** | Hospital length-of-stay for treated (alteplase or EVT) stroke patients (reduce) [adjusted for age, sex and stroke severity to account for imbalance between groups] | | - date of discharge from hospital (including in-patient rehabilitation)<br>- date of admission |
| **Secondary** | Door-In-Door-Out time | Reduce the median door-in-door-out time for transfers for EVT to a median of 50 min | - time of departure from first hospital<br>- time of arrival at first hospital |

| Hierarchy | Measure | Goal | Source measures |
|---|---|---|---|
| **Secondary** | Door-to-Groin-Puncture | Reduce the median door-to-groin-puncture time for all patients treated with EVT to a median of 60 minutes | - time of arrival at EVT capable hospital<br>- time of groin puncture |
| **Secondary** | First-Medical-Contact to Needle | Reduce time to alteplase treatment | - time of 911 call<br>- time of alteplase start |
| **Secondary** | First-Medical-Contact to Groin Puncture | Reduce time to EVT treatment | - time of 911 call<br>- time of groin puncture |

**Quality Improvement Intervention:**
The Institute for Healthcare Improvement's (IHI) Breakthrough Series Collaborative model (herewith referred to as the *Improvement Collaborative*)[26] will be used to implement improvement of acute stroke treatment across Atlantic Canada. This intervention has been used to improve and implement evidence-based best practice, though with mixed results.[27-31] The Improvement Collaborative is derived from the "Model for Improvement" utilizing the PDSA (Plan-Do-Study-Act) cycles, which was developed within the industrial engineering community.[32-35] However, industrial engineers have rarely been included in Improvement Collaboratives in most health care settings.

Our interdisciplinary research team includes experts from stroke neurology, industrial engineering, emergency medicine, interventional neuroradiology, emergency medical services, and implementation science in the execution of the Improvement Collaborative intervention. This approach will be facilitated by an established relationship between industrial engineering and the health care systems in Atlantic Canada. The Improvement Collaborative intervention has been used successfully for the improvement of acute stroke treatment in Alberta; a project led by the N. Kamal (NPI).[36] It successfully reduced door-to-needle times across Alberta to 36 minutes from 68 minutes;[37,38] Alberta was the first jurisdiction in the world to achieve this level of improvement across an entire population. However, it has not been used to improve access to stroke treatments, or to improve efficiency of EVT treatment. Additionally, this intervention was only applied to a single health system within one province. Although the focus of the Alberta initiative was only on improving thrombolysis efficiency, the initiative increased the percent of patients that received thrombolysis to 15.7% from 9.3%, and an additional 13% of patients that received thrombolysis were able to return home from acute care.[39] Based on the experiences in Alberta,[36] the Improvement Collaborative intervention will be modified to be more effective and efficient.

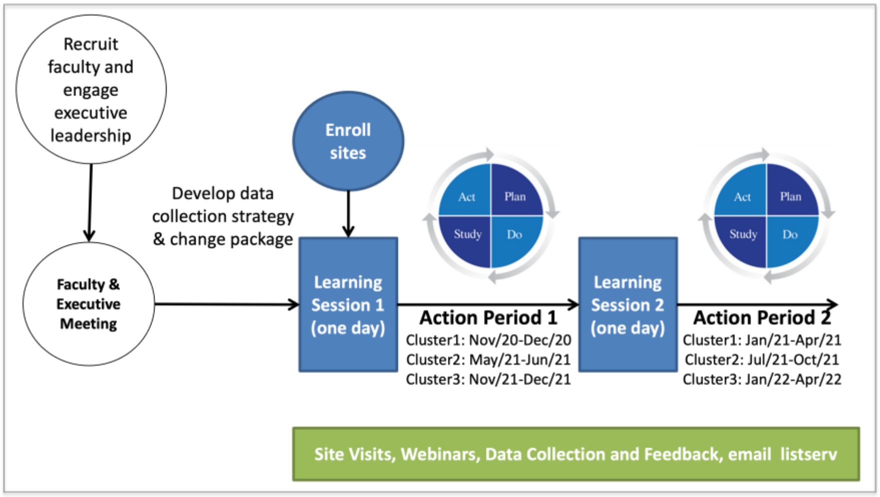

*Figure 1. Improvement Collaborative Intervention*

Figure 1 shows the overall Improvement Collaborative intervention that will be used for this project.

Prior to the commencement of the improvement Intervention, recruitment and commitment is needed at the provincial level for each of the provinces to ensure engagement of key local clinical experts and executive leadership. The research team is a microcosm of the implementation environment, with clinical and administrative leaders for stroke care in each province. A face-to-face meeting will be facilitated with this research team, research collaborators and additional key individuals that are later identified (shown in the white circles). This face-to-face meeting will be done in addition to regular weekly meetings. The measurement strategy and data collection mechanisms will then be finalized prior to the start of the first Collaborative (see *data collection* sub-section below).

One month prior to the commencement of the improvement intervention for each cluster (see *Stepped Wedge Trial* section below), all stroke-treating hospitals will be formally enrolled. The process will include assembling an interdisciplinary team of engaged individuals at each site. Teams from each stroke site (hospital) will include at least one physician (emergency physician, radiologist, neurologist, or other stroke physician), stroke program coordinator, ED nurse, paramedic lead (e.g. regional supervisor), diagnostic imaging representative, and an administrator. Additional disciplines can be included, as deemed necessary by site teams. Executive leadership must sign off on the team's participation in the Improvement Collaborative, and preferably be active participants.

All enrolled sites will cycle between alternating Learning Sessions and Action Periods. During the Learning Session, site teams will travel to a central location for a one-day workshop. The workshop will provide presentations on the evidence underlying the improvement effort and the rationale for a call-to-action; they will hear from peer teams about how access and efficiency can be improved; additionally, each hospital's improvement team will plan their own changes. Sample agendas for the Learning Sessions are included in the Appendix A. During the Action Period, the sites will test the changes that were discussed and planned during the Learning Sessions. Sites will customize these new processes for their context using the Model-for-Improvement's PDSA cycles. Supports will be available to the sites throughout the 6-month process with site visits (agenda provided in Appendix B), data audit and feedback (letter sample provided in Appendix C), webinars (curriculum provided in Appendix D), as well as email discussion through a listserv provided by the research team.

The ideal changes that will be provided in the Change Package and Learning Session presentations are based on existing research studies (Figure 2).[40] Changes will be customized by the sites for their context during the Action Periods. The industrial engineering researchers and trainees, who are part of the research team, will work locally with each site during the Action Periods using techniques such as simulation modeling to test changes, evaluate scenarios, and provide proof of concept. As shown in Figure 2, ambulance transport to the first hospital is critical in reducing treatment delays, as they play a pivotal role in pre-notification, pre-identification and pre-registration of the incoming patients, which allows the stroke team to lookup the patients history, medication and contraindication. Additionally, Emergency Medical

Services (EMS) including air transport plays a vital role in developing centralized and standardized processes to reduce transport delays that ensure that the patient is transported to the right hospital, and to reduce transfer delays for patients being transferred for EVT. Therefore, the research team includes representatives from Emergency Medical Services (EMS) including Critical Care Transport (CCT) services which include ground, fixed-wing and helicopter transport modalities. Additional details about this is provided in the *Feasibility* section below.

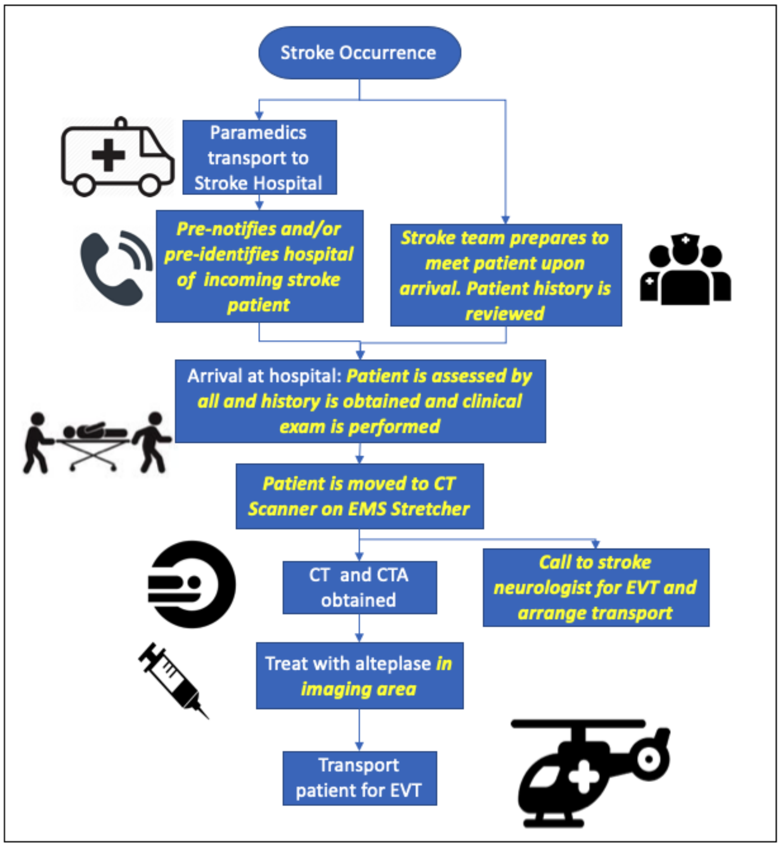


*Figure 2. High-level ideal acute stroke treatment process. Key changes are shown in yellow.*

**Data Collection:**

Critical to evaluating the impact of the intervention is a robust data collection system. The research team has significant experience and access to acute stroke data collection systems. QuICR Alberta Stroke Program developed a clinical registry to capture data for all patients that were treated with alteplase and/or EVT in Alberta (https://ucalgary.ca/quicr/registry). This registry was developed by N Kamal (NPI) and MD Hill (co-applicant). Nova Scotia has also created robust data collections systems. The Cardiovascular Health Nova Scotia Stroke (CVHNS) Registry includes population-level data that includes treatment data including treatment efficiency. This registry is the responsibility of N Gill at NSHA's (Nova Scotia Health Authority) CVHNS program, who is a partner on this grant. Additionally, the QEII maintains an Acute Stroke Registry,[41] developed by S Phillips (co-applicant). It is a prospective stroke registry that includes information about Acute Stroke Protocol activations and recanalization therapy at the QEII. PEI, NB, and NL have also been active in collecting data on acute stroke treatment for their population. All provinces have relevant treatment data that is not already captured in administrative datasets. By combining our expertise with the QuICR registry, the CVHNS Stroke Registry and the QEII Acute Stroke Registry, we will develop a robust data collection system capable of capturing information on stroke patients and treatment. There is potential for the expansion of the QuICR registry to some or all of the Atlantic provinces with due consideration of privacy requirements within each province. The possibility of having a heterogenous data collection system based on each province's need will also be explored, as we want to ensure that the process is streamlined, useful, and sustainable.

In Newfoundland and Labrador, the data collection will be done through a provincial stroke coordinator in Newfoundland and Labrador, who has provincial oversight. The stroke coordinator will periodically access charts and manually extract data. This data will be de-identified by the stroke coordinator, and the data will then be securely transferred to the ACTEAST Principal Investigator.

**Stepped Wedge Trial (SWT):**

The quality improvement intervention described above will be implemented within a Stepped-Wedge Trial (SWT) to measure its effectiveness. In health and medicine, the randomized controlled trial (RCT) design is widely accepted as the gold standard for evaluating the efficacy of an intervention on patient outcomes. However, in quality improvement and implementation interventions, it is not possible to randomize at the patient level since changes are made at the system level. For this reason, a cluster trial design is increasingly being proposed,[42] where an entire site is randomized to receive or not receive the intervention. However, there are several ethical issues with this design, as patients treated at hospitals in the control arm never receive the intervention. The SWT design alleviates these concerns by introducing the intervention to all participating hospitals in a sequential step-wise approach.[43]

| Sites | Lead in Phase (May/20-Oct/20) | Phase 1 (Nov/20-Apr/21) | Phase 2 (May/21-Oct21) | Phase 3 (Nov/21-Apr/22) | Phase 4 (May/22-Oct/22) |
|---|---|---|---|---|---|
| **Cluster 1** [NS; n=10] | Retrospective Data Collection | Intervention | Yes | Yes | Yes |
| **Cluster 2** [NB,PEI; n=12] | Retrospective Data Collection | No | Intervention | Yes | Yes |
| **Cluster 3** [NL; n=12] | Retrospective Data Collection | No | No | Intervention | Yes |

*Figure 3. Stepped Wedge Trial Design*

The evaluation of this *improvement intervention* will be conducted through SWT (Figure 3). In the SWT, all sites will be assigned to a group or cluster. Each cluster will go through the intervention at different times. Prior to going through the intervention, all clusters are in the control phase, while after the intervention, all clusters will have the intervention fully implemented. The intervention will be 6 months in length. In Figure 3, the orange areas show the "control" periods where the intervention has not yet started and the green areas show the periods after intervention has been completed. The data collected in the orange and the green phases will be analysed, and data during the implementation phase will be excluded.

All Learning Sessions and other aspects of the intervention will be delivered in English. There will be considerations provided for Cluster 2 (New Brunswick and PEI) for the French speakers. The Learning Sessions in Cluster 2 will be delivered in English because there is no budget for simultaneous; however, all powerpoint presentations will be available in French in both paper and electronic copies. All recruitment material will be provided in both languages for Cluster 2.

*Sample Size Calculation:* The proposed trial is quasi-randomised insofar as the sites will not be randomized to a cluster for pragmatic reasons. The data for this trial includes all ischemic stroke patients in the entire province regardless of the number of sites that formally enrolled. The trial is therefore a population based study; no sample size was calculated. However, we did calculate the sample size required to reach significance for out objectives.

We assume that the primary outcome of improvement is the proportion patients receiving treatment, and we assume that the sample size calculation is based upon a cluster design. With 3 clusters, 10 or more sites per cluster, an estimated 10 patients per site during each study period, an intra-cluster correlation of 0.8, conventional alpha of 0.05, the trial will have a power of 82% to detect the desired 5% improvement in the proportion of patients receiving treatment. The same design will have 96% power to detect a 10-minute reduction in mean treatment time with estimated common standard deviation of 30 minutes assuming an intra-cluster correlation of 0.3.

*Statistical Analysis:*

All missing values for age, sex, and NIHSS was imputed before running the adjusted analysis; using the K-Nearest Neighbour algorithm (k-NN), a multi variate approach that identifies k nearest neighbours for a missing data point from all complete instances in a dataset by calculating the Euclidean distance between them.

For the comparison analysis related to all ischemic stroke patients, unadjusted proportion comparison is conducted using $\chi^2$ analysis, while a mixed effects logistic regression with cluster as random effects variable is used for adjusted analysis. For analysis related to all treated patients, unadjusted analysis comparison is conducted using $\chi^2$ for proportions and Wilcoxon rank-sum test for continuous variables. The comparison analysis uses a mixed effects logistic regression for binary/categorical variables and a mixed effects quantile regression for continuous variables. In these analyses, age, sex, and NIHSS are fixed effects variables while cluster was a random effects variable. We did not include the time of the intervention as cluster captures the temporality of the stepped-wedge trial, and the overall intervention period is short. When analyzing results by cluster, we used a simple quantile regression model.

*Consideration of Sex and Gender:* The final analysis will determine if the same level of improvement was achieved for female and male patients, as there is evidence that women are treated for acute stroke less than men.[44-46] Differences in treatment proportions by sex will be compared between the orange and green phases (Figure 3). This analysis will be exploratory, but will generate additional hypotheses, so that we can better understand the difference in treatment of acute stroke patients between women and men. Gender differences will be explored in the qualitative evaluation. Further studies can then use this data for enhanced understanding of sex and gender differences.

*The work plan and timeline* for this project are shown in the table below.

| Workplan | Timeline |
|---|---|
| **Obtain ethics approval, and create data collection system** | Mar/20-Sep/20 |
| **Cluster 1 intervention: improvement collaborative (NS)** | Nov/20-Apr/21 |
| **Cluster 2 intervention: improvement collaborative (NB and PEI)** | May/21-Oct/21 |
| **Cluster 3 intervention: improvement collaborative (NL)** | Nov/21-Apr/22 |
| **Wrap-up activities including the finalization of all data collection** | May/22-Feb/23 |
| **Data Analysis and dissemination** | Mar/23-Feb/24 |

Note: the site enrolment prior to the intervention will occur in parallel with the previous activity.

**Qualitative Evaluation:**

A qualitative evaluation of the intervention will also be conducted using a theory-driven, evidence-based approach to understand how the *improvement intervention* was implemented, and to identify key implementation barriers, facilitators or other influences on implementation. This knowledge can be used to both understand and modify implementation strategies, and their potential scale. We will investigate provider perceptions of the *improvement intervention* by provider type (e.g. physicians, nurses, administrators, paramedics etc.) and explore perceptions of specific intervention components that were most effective such as the Learning Sessions, site visits, webinars and data feedback. This interpretive, formative evaluation[47] will provide a working hypothesis of the barriers, facilitators and other contextual influences explaining the improvement collaborative. Face-to-face semi-structured interviews will be conducted after the collaborative. In addition to the face-to-face interviews, participants in the Improvement intervention will be asked to complete an online questionnaire prior to attending the first Learning Session after their site has been enrolled, and the participant has been listed as a team member. For Cluster 2 (New Brunswick and PEI), the online questionnaire will be available in French. Participation in the interview after the intervention will be voluntary. For Cluster 2 (New Brunswick and PEI), paticipants will be given the option to participate in the interview in either French or English. ACTEAST will hire a bilingual research assistant for a period of 2 months to complete the interviews for this cluster.

The interviews and focus groups will be conducted by the ACTEAST Principal Investigator or an ACTEAST research assistant who may be hired at a later time.

Based on previous barrier-facilitator mapping in Alberta, we will apply a modified deductive coding framework built from the combined domains of the Consolidated Framework for Implementation Research (CFIR)[48] and the Theoretical Domains Framework (TDF).[49] This coding framework will be applied specifically with the intent to view and assess the feedback obtained from providers through an implementation lens. Line-by-line coding of transcripts will identify excerpts that report barriers or facilitators to the implementation, and then coded to align with TDF and/or CFIR categories. This initial coding will be followed by a domain-by-domain review of each code category for data fidelity. High frequency codes will be explored to identify potential sub-themes using thematic analysis.[50]

Domain codes will be summarized by frequency and distribution across the combined modified CFIR and TDF domains. Content for the most frequent domains arising in the deductive analysis will be detailed using narrative summaries. We will then pivot the coded data and analyze interviews both by province, site, and site location (urban/rural) to examine how sites differ in their perception of barriers and facilitators. Additional details are provided Appendix E.

**Feasibility:**
There are many challenges with carrying out this project across four provinces in Canada. Developing relationships with the appropriate administrators and clinicians is critical. ACTEAST has assembled the right team to ensure success of the project. The research team includes:

*N. Kamal:* The Nominated Principal Applicant is an engineer and Assistant Professor at Dalhousie University's Department of Industrial Engineering with 7 years of experience working on improving stroke care in both Alberta and British Columbia, where she used the *Improvement Collaborative* methodology extensively. She has done extensive research on the causes of delays in stroke treatment[51] as well as the changes that lead to faster treatment.[40] Her experience in improving stroke treatment in Alberta will be incorporated into ACTEAST.

*Stroke Neurologists (Halifax):* Stephen Phillips and Gordon Gubitz are stroke neurologists at QEII in Halifax and full Professors at Dalhousie University. Their existing relationships with Nova Scotia's health system and their knowledge of the availability of acute stroke data will provide valuable foundation and a critical path to success for this research.

*Clinical Trial Leadership:* Michael Hill and Bijoy Menon are stroke neurologists at Foothills Medical Centre (Calgary, AB), and they hold academic appointments at the University of Calgary. They are both experts in the design of clinical trials. Dr. Hill led the ESCAPE trial, which was one of the foremost RCTs that proved the efficacy of EVT.[4] Dr. Hill is also a leading expert in trial design including pragmatic clinical trials such as the SWT design. Dr. Menon is a trained epidemiologist and trial methodologist and brings expertise in the design and conduct of pragmatic and registry embedded clinical trials. They will help with the design and implementation of the SWT for ACTEAST and in planning and implementing all analysis.

*Industrial/Systems Engineers:* John Blake and Peter Vanberkel are Industrial Engineers and Associate Professors at Dalhousie University. Dr. Blake's research specializes in simulation and optimization in health care. For this project, he will provide leadership to hospital teams and trainees on how simulation can be used to test changes. Dr. Vanberkel has extensive experience in optimizing the ambulance service in Nova Scotia, which will be useful to optimize transfer protocols within Nova Scotia as well as between other Atlantic provinces.

*Neurointerventional Radiologists:* David Volders and Thien Huynh are neurointerventional radiologists at the QEII Health Sciences Centre in Halifax, NS. They hold academic appointments at Dalhousie University. They will provide critical insight and action into improving the efficiency of EVT treatment across Atlantic Canada.

*Emergency Medical Services:* Alix Carter, Judah Goldstein and Patrick Fok are clinical and research experts for EMS in NS with connections with EMS in NB and PEI. Dr. Carter is the Medical Director of research for EMS in NS; Dr. Judah Goldstien is the Research Coordinator as well as a paramedic; Dr. Patrick Fok is a medical oversight Physician for Lifeflight in NS (air ambulance). All three hold academic appointments with Dalhousie University. They will provide critical linkages with EMS across NS, and assist with streamlining existing protocols to improve initial transport and transfer. They also have linkages with EMS in NB and PEI through the umbrella organization, Medavie.

*Medical Leadership in PEI and NL:* Heather Williams is a neurologist in PEI and the clinical lead for stroke in the province. She will provide clinical leadership in PEI. E. Van Der Linde, G. Browne, and B. Metcalfe are 3 physicians on our research team from NL. Van Der Linde is an emergency physician and stroke champion in Clarenville, NL. He will represent the emergency physician group. Dr. Greg Browne is a vascular surgeon in St. John's NL, and he has been working to set-up EVT services in NL. Dr. Brian Metcalfe is the Medical Chief for EMS in NL, and he will assist with streamlining transport and transfer of stroke patients across NL.

***Health Care Administration Collaboration:*** ACTEAST also has representation from the key administrators responsible for stroke care across all four Atlantic provinces. The following people are collaborators on this project, and they will provide linkages to stroke hospitals in their province or health authority, as well as linkages to key clinical leadership and EMS: 1) in NS, Neala Gill is the Program Manager for the NSHA's Cardiovascular Health Nova Scotia (CVHNS); 2) in PEI, Carolyn MacPhail and Trish Helm-Neima are the Manager of Chronic Disease Prevention and Management and Provincial Stroke Coordinator (respectively) for Health PEI; 3) in NL, Cassie Chisholm is the Director of Primary Health Care for the Government of Newfoundland & Labrador (department responsible for stroke); 4) Provincially in NB, Noortje Kunnen works for the Government of NB and she is responsible for stroke care; 5) in Horizon Health Network in NB, Nicole Tupper is the Executive Director responsible for stroke; and 6) in Vitalité Health Network in NB, Nadia D'Astous is the stroke coordinator; and 7) for EMS in NB, Edgar Goulette is the Vice President of Quality, Patient Safety and Education at Ambulance New Brunswick who will support EMS engagement for NB.

In addition to the research team and provincial partners, we have additional collaborators that will contribute to the success for this project. Scott Theriault is a stroke survivor, who was treated with EVT at the QEII. He will represent the patient on our research team, and provide the patient voice throughout our improvement intervention. K. Mrklas is a Knowledge Translation Implementation Scientist at Alberta Health Services, and is a national leader on knowledge translation and will ensure we are following best practices in the incorporation of implementation science. P. Lindsay is Director of Stroke at the Heart and Stroke Foundation, and she has connections with stroke clinicians and administrators from across Canada.

**Knowledge Translation and Impact:**
The ACTEAST project was conceptualized, designed and will be executed using an integrated Knowledge Translation (IKT) approach. We have deliberately engaged representative knowledge users, including patients, from all 4 provinces on the team to develop this grant, and to act as local brokers within their respective settings. We have engaged clinical and administrative decision makers to ensure oversight and decision making is locally aligned.

If successful, this IKT project will have significant impact across Atlantic Canada. ACTEAST has the potential to develop referral patterns and processes that will improve access and efficiency of treatment for acute stroke patients across Atlantic Canada. The health impacts of ACTEAST includes both *Health Status* (less disability) and *Determinants of Health* (efficiency, effectiveness, and appropriateness).[52] The estimated benefit is an additional 7-15% of ischemic stroke patients will gain functional independence. This increase translates to *260 to 550 more patients every year will be able to return home* after their stroke in Atlantic Canada.[3,5] These benefits will be especially apparent in rural and remote communities, which are presently underserved. Additionally, stroke is a very expensive disease, as patients often require lengthy stays in hospital for rehabilitation, which will be reduced through the objectives of the ACTEAST project. If more patients can access efficient treatment, there will be significant cost avoidance to the health system across Atlantic Canada. Based on studies estimating the cost of treatment by stroke outcomes,[53] approximately *$7.8 million per year in health care costs will be avoided* across Atlantic Canada, based on an additional 260 patients being able to return home with no disability. This significant potential rate of return on a relatively small investment in a short time frame is unsurpassed by any treatment that is currently available to patients.[9]

The challenges of less than optimal access and efficiency of treatment is not unique to Atlantic Canada. The successful implementation of evidence-based treatment with both alteplase and EVT for ischemic stroke as a coupled strategy across health systems can have significant clinical benefit for those afflicted by this disease. The dissemination strategy is provided in Appendix F.

**Budget:**

This is a 3-year project. The annual budget for this project is as follows:

| | YEAR 1 | | | | | |
|---|---|---|---|---|---|---|
| Item | Number | Cost per unit | Total | Matching Source | Matching amount | Amount from CIHR |
| Faculty Travel | 5 | $700 | $3,500 | | | $3,500 |
| Participant Travel | 100 | $300 | $30,000 | CVHNS | $20,000 | $10,000 |
| Learning Sessions | 2 | $4,000 | $8,000 | QuICR Grant | $8,000 | $0 |
| Trainees | 2 | $18,000 | $36,000 | | | $36,000 |
| Site Visit | 12 | $800 | $9,600 | | | $9,600 |
| Dissemination | 2 | $1,800 | $3,600 | | | $3,600 |
| CME Accreditation | 2 | $475 | $950 | | | $950 |

| Item | Number | Cost per unit | Total | Matching Source | Matching amount | Amount from CIHR |
|---|---|---|---|---|---|---|
| Administrative Support | 0.4 | $40,000 | $16,000 | | | $16,000 |
| Data Collection System | 1 | $15,000 | $15,000 | | | $15,000 |
| **Total** | | | **$122,650** | | **$28,000** | **$94,650** |

| | **YEAR 2** | | | | | |
|---|---|---|---|---|---|---|
| **Item** | **Number** | **Cost per unit** | **Total** | **Matching Source** | **Matching amount** | **Amount from CIHR** |
| Faculty Travel | 6 | $700 | $4,200 | | | $4,200 |
| Participant Travel | 100 | $300 | $30,000 | | | $30,000 |
| Learning Sessions | 2 | $4,000 | $8,000 | | | $8,000 |
| Trainees | 2 | $18,000 | $36,000 | | | $36,000 |
| Site Visit | 12 | $800 | $9,600 | | | $9,600 |
| Dissemination | 2 | $1,800 | $3,600 | QuICR Grant | $3,000 | $600 |
| CME Accreditation | 3 | $475 | $1,425 | | | $1,425 |
| Administrative Support | 0.6 | $40,000 | $24,000 | | | $24,000 |
| Data Collection system | 1 | $5,000 | $5,000 | | | $5,000 |
| **Total** | | | **$121,825** | | **$3,000** | **$118,825** |

| | **YEAR 3** | | | | | |
|---|---|---|---|---|---|---|
| **Item** | **Number** | **Cost per unit** | **Total** | **Matching Source** | **Matching amount** | **Amount from CIHR** |
| Faculty Travel | 6 | $700 | $4,200 | | | $4,200 |
| Participant Travel | 100 | $300 | $30,000 | | | $30,000 |
| Learning Sessions | 2 | $4,000 | $8,000 | | | $8,000 |
| Trainees | 2 | $18,000 | $36,000 | Dalhousie Start-up | $15,000 | $21,000 |
| Site Visit | 12 | $800 | $9,600 | | | $9,600 |
| Dissemination | 2 | $1,800 | $3,600 | QuICR Grant | $3,000 | $600 |
| CME Accreditation | 3 | $475 | $1,425 | | | $1,425 |
| Administrative support | 0.6 | $40,000 | $24,000 | | | $24,000 |
| Data Collection system | 1 | $5,000 | $5,000 | | | $5,000 |
| **Total** | | | **$121,825** | | **$18,000** | **$103,825** |

*Faculty Travel:* These funds allow investigators to travel to the Learning Sessions. It includes mileage and 1-2 nights accommodations.

*Participant Travel:* These funds allow the participants to travel to the Learning Sessions. It includes 1-2 nights accommodations and mileage.

*Learning Sessions:* These funds include cost to rent the room to hold the Learning Sessions, catering for all the attendees, and AV costs.

*Trainees:* These funds are for 2 masters or PhD students, who are working on research related to ACTEAS.

*Site Visits:* These funds will cover travel costs for 1-2 investigators to visit each site during the Improvement Collaborative.

*Dissemination:* These funds cover travel costs to conferences to present the results from this study, and publication costs.

*CME Accreditation:* These funds allow the Learning Session to be accredited for continuing medical education credits.

*Administration Support:* These funds cover the costs of a part-time coordinator.

*Data Collection System:* These funds cover the costs of a data collection system.

### Appendix A: Learning Session Agendas

#### ACTEAST Enhancing Acute Stroke Treatment Improvement Collaborative

#### Learning Session 1

| Time | Topic | Presenter |
|---|---|---|
| **9:00am – 9:15am** | Introductions and ACTEAST Overview | Noreen Kamal, PhD |
| **9:15am – 9:40am** | Stroke Patient Story: Treatment with EVT | Scott Theriault |
| **9:40am – 10:10am** | Acute Stroke Treatment: Efficacy and Importance of Efficiency<br>*7 min question and answer* | Dr. Stephen Phillips |
| **10:10am – 10:40am** | The Alberta Experience with Improving DTN times and Patient Outcomes<br>*7 min question and answer* | Noreen Kamal, PhD |
| **10:40am – 11:00am** | *Coffee/Tea/Health Break & Networking* | |
| **11:00am – 11:20am** | Changes that Improve Efficiency for Acute Stroke Treatment<br>*7 min question and answer* | Noreen Kamal, PhD & Dr. Gord Gubitz |
| **11:20am – 11:35am** | Importance of conducting a CTA immediately after the CT<br>*7 min question and answer* | Dr. David Volder & Dr. Jens Heidenreich |
| **11:35am – 12:00pm** | EMS and Lifeflight: Understanding Provincial Transfer<br>*7 min question and answer* | Dr. Patrick Fok & Judah Goldstein, PhD |
| **12:00pm – 12:15pm** | Improving Efficiency in Hospital X<br>*5 min question and answer* | Hospital X representative |
| **12:15pm – 1:15pm** | *Lunch & Networking* | |
| **1:15pm – 1:45pm** | Process Mapping<br>*7 min question and answer* | Noreen Kamal, PhD & John Blake, PhD |
| **1:45pm – 2:30pm** | *Sites map their process* | Small Groups |
| **2:30pm – 2:45pm** | *Coffee/Tea/Health Break & Networking* | |
| **2:45pm – 3:00pm** | Testing Changes: PDSA cycles<br>*5 min question and answer* | Noreen Kamal, PhD & Peter Vanberkel, PhD |
| **3:00pm – 3:45pm** | *Planning improvement with your team* | Small Groups |
| **3:45pm – 4:15pm** | *Report back* | All |
| **4:15pm – 4:30pm** | Wrap-up, Final Thoughts, and Next Steps | Noreen Kamal, PhD & All |

### ACTEAST Enhancing Acute Stroke Treatment Improvement Collaborative

#### Learning Session 2

| Time | Topic | Presenter |
|---|---|---|
| **9:00am – 9:15am** | Introductions | Noreen Kamal, PhD |
| **9:15am – 9:45am** | How far have we come? Review of data<br>*7 min question and answer* | Noreen Kamal, PhD & Dr. Stephen Phillips |
| **9:45am – 10:15am** | Fastest Site in the Province<br>*7 min question and answer* | Hospital Representative |
| **10:15am – 10:30am** | *Coffee/Tea/Health Break & Networking* | |
| **10:30am – 10:50am** | Pre-Notification/Pre-Identification/Pre-Registration<br>*5 min question and answer* | Dr. Gordon Gubitz |
| **10:50am – 11:10am** | Direct to CT/Swarm/Quick Neuro exam<br>*5 min question and answer* | TBD [a site that has implemented it] |
| **11:10am – 11:30am** | Giving Alteplase in the CT Scanner or Imaging area - *5 min question and answer* | TBD [a site that has implemented it] |
| **11:30am – 11:50am** | Creating a alteplase/stroke kit<br>*5 min question and answer* | TBD [a site that has implemented it] |
| **11:50am – 12:10pm** | Early notification to EMS for Transfer for EVT<br>*5 min question and answer* | TBD [a site that has implemented it] |
| **12:10pm – 1:10pm** | *Lunch & Networking* | |
| **1:10pm – 2:00pm** | Facilitated Discussion: Breaking down Facilitators and Barriers to Improvements | All |
| **2:00pm – 2:15pm** | *Coffee/Tea/Health Break & Networking* | |
| **2:15pm – 3:00pm** | *Planning improvements with your team* | Small Groups |
| **3:00pm – 3:45pm** | *Report Back* | All |
| **3:45pm – 4:00pm** | Final Thoughts, Next Steps | Noreen Kamal, PhD |

### APPENDIX B: Site Visit Agenda

Invitation to anyone involved with Acute Ischemic Stroke patient treatment at this hospital

| Topic | Presenter | Time Allotment |
|---|---|---|
| **Introductions** | Noreen Kamal | 15 minutes |
| **Stroke Treatment** | Physician lead (Dr. Phillips) | 30 minutes |
| **ACTEAST Overview and Site's Progress** | Noreen Kamal | 30 minutes |
| **Site Presentation** | Site representative | 45 minutes |
| **Discussion about progress and next steps** | All | 1 hour |

The formal meeting can be followed by additional activities such a site tour.

### Appendix C: Data Feedback Letter Example

Dear <Site X Team>,

Thank you for your participation in the ACTEAST Improvement Collaborative. This initiative is aiming to improve the proportion of ischemic stroke patients that receive treatment, and the DTN times for patients treated with alteplase, as well as the door-in-door-out times for patients transferred for EVT.

Please find attached your DTN data for <month year>. The first chart shows the DTN times for each patient treated with tPA across the province. The site and DTN times are labeled, and the zones are separated by colour. The median DTN zin for the province was 31.5 minutes with 81% of all patients treated in 60 minutes or less.

Your site treated 2 patients this month out of a total of 12 ischemic stroke patients (16.7%), which is very good. Your DTN times for these patients were 77 and 78 minutes, which is well above our target of 30 minutes. Let’s work to bring down, please feel free to contact me to discuss further. Your site also transferred one of these patients for EVT with Door-in-Door-Out (DIDO) time of 100 minutes The new provincial standard for treatment is a median DTN time of 30 minutes or less and a median DIDO time of 50 minutes, which I know that you are capable of achieving and setting a strong example to the rest of the province.

If you need assistance implementing changes at your site, please do not hesitate to contact Noreen Kamal at QuICR, via or via telephone 902-494-3293.

Sincerely,

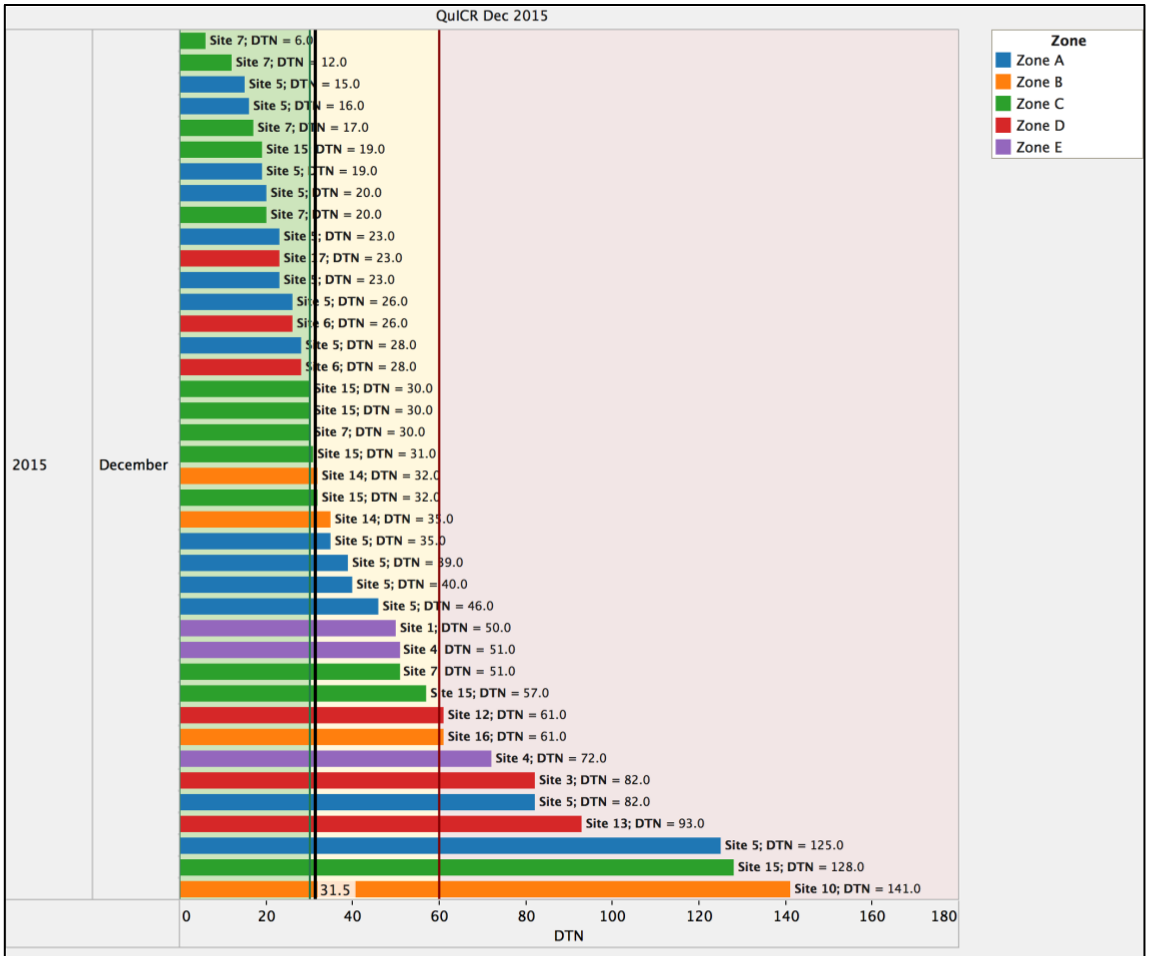


Door-to-Needle times for Alberta in December 2016. Median is shown by verticle line (31.0 minutes).

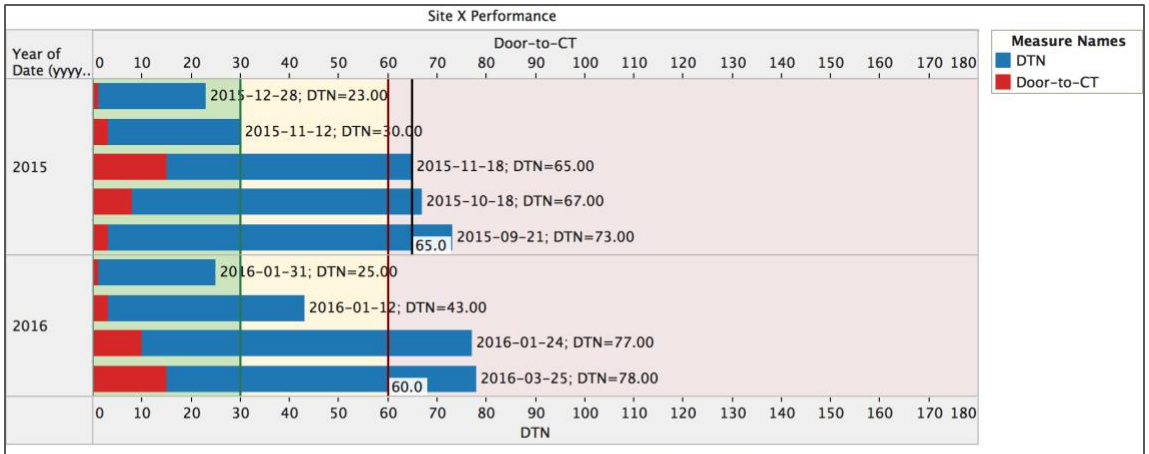


Door to Needle times and Door to CT times for XYZ Hospital since the start of the QuICR DTN initiative. Median is shown by verticle line for each year.

### Appendix D: Webinar Curriculum

| Webinar Topic | Description |
|---|---|
| **Implementing Changes to Improve the process** | A review of the change package that lists the specific changes to improvement treatment process including examples of how it has been implemented at specific sites. The webinar will include a discussion with participants |
| **Understanding Contrast Risk for CTA** | A review of the evidence of the risk of contrast on kidneys when conducting a CTA. Webinar given by clinical expert |
| **Provincial EMS** | A provincial EMS representative will present on the stroke transport process and EVT transfer process. Participants will have an opportunity to ask about issues that they have been experiencing with EMS followed by discussions on how these issues can be resolved |
| **Case Review 1** | A site will present 2 cases: one that went well with fast treatment times and/or transfer times, and another one that did not go well with much slower treatment times. |
| **Case Review 2** | Another site will present 2 cases: one that went well with fast treatment times and/or transfer times, and another one that did not go well with much slower treatment times. |
